# Factors associated with lower COVID-19 vaccine uptake among populations with a migration background in the Netherlands

**DOI:** 10.1101/2024.11.07.24316886

**Authors:** Bente Smagge, Lisanne Labuschagne, Joyce Pijpers, Annika van Roon, Susan van den Hof, Susan Hahné, Hester de Melker

**Affiliations:** Centre for Infectious Disease Control, National Institute for Public Health and the Environment (RIVM), Bilthoven, the Netherlands

## Abstract

**Background:** In high income countries, the incidence of severe COVID-19 has been disproportionally high among persons with a migration background. We examined determinants of being unvaccinated against COVID-19 in the Netherlands among four large populations of non-Dutch origin with below average vaccination coverage.

**Methods:** A nationwide study of determinants of being unvaccinated in the 2021 primary COVID-19 vaccination round in adults and 2022 autumn booster in those ≥60 years was performed within the Netherlands’ populations of Dutch-Caribbean, Moroccan, Surinamese and Turkish origin. Using vaccination registry and individual and household level sociodemographic and socioeconomic data, we examined the association between each potential determinant and being unvaccinated using multivariable logistic regression. In addition, we computed population attributable fractions (PAFs).

**Results:** Among these populations of non-Dutch origin, the odds of being unvaccinated in both the primary vaccination round and the 2022 booster round were higher among younger persons, migrants with two foreign-born parents, inhabitants of highly or extremely urban areas and persons with low medical risk, lower income and lower education level. The higher odds of non-uptake for migrants with two foreign-born parents (reference: Netherlands-born child with one foreign-born parent) weakened but persisted after adjusting for socioeconomic variables in the populations of Dutch-Caribbean, Moroccan and Surinamese origin (Dutch-Caribbean: aOR_primary_=3.39 vs. 2.51, aOR_booster_=2.51 vs. 1.99, Moroccan: aOR_primary_=2.16 vs. 1.80, Surinamese: aOR_primary_=1.21 vs. 1.09, aOR_booster_=2.22 vs. 1.99), and inversed in the population of Turkish origin (aOR_primary_=1.10 vs. 0.93), while adjusting for additional variables had little effect on the estimate. Similarly to the aORs, the PAFs of young age, being a migrant with two foreign-born parents and having low income, low education level and low medical risk were highest.

**Conclusion:** Age, urbanisation level, medical risk, income, education level and migration background were associated with COVID-19 vaccination status among populations of non-Dutch origin. Socioeconomic status only partially mediated the effect of migration background. Although these findings provide some guidance to target vaccination programmes, qualitative and survey-based research is needed to further understand reasons behind lower vaccine uptake and design (community-based) interventions to improve health equity.

## 1. Introduction

The COVID-19 pandemic increased health inequities both between and within countries. Not only are older adults and individuals with underlying medical conditions at higher risk of severe COVID-19, also socioeconomically disadvantaged groups and ethnic minorities and migrants have been disproportionally affected [1, 2]. Having a migration background has been associated with increased risk of infection and severe disease in several high-income countries [1, 3-5]. Also after the initial waves of the pandemic, COVID-19 hospitalisation rates have remained higher in these populations [6]. COVID-19 vaccination offers protection against severe disease and death and booster rounds have been rolled out to improve and maintain protection over time [7]. To reduce disparities, vaccination coverage should be homogeneously high. However, vaccine uptake is reported to be below average among subpopulations, including migrant and ethnic minority populations [2, 8-11].

In the Netherlands, vaccination against COVID-19 started on 6 January 2021 [7]. The primary vaccination series was implemented in a step-wise manner, until the whole population aged 12 years and older had been invited by mid-July 2021. While the primary series remained available, successive booster rounds started in November 2021, February 2022 and every autumn since 2022 [12]. Uptake of at least one dose of COVID-19 vaccine and vaccine uptake during the 2022 autumn booster campaign was generally lower among individuals of non-Dutch compared to Dutch origin [2, 11]. In the Dutch National Immunisation Programme differences in vaccine uptake by (parents’) country of origin have also been observed [13-16].

Previous registry-based research into sociodemographic determinants of COVID-19 vaccine uptake in the overall Dutch population have identified age, income, socioeconomic position, whether a person or their parent(s) were born in the Netherlands and political preference as important factors [2, 11]. The current study seeks to explore determinants of being unvaccinated against COVID-19 in the primary vaccination round and in the 2022 autumn booster round among four populations of non-Dutch origin.

## 2. Methods

### 2.1. Study design and population

We performed a retrospective database study. For the analyses of the primary vaccination series, we included the entire population aged 18 years and older as registered in the Personal Records Database of the Netherlands on 5 January 2021. For the analyses of the 2022 autumn booster campaign, the study population comprised all individuals aged 60 years and older registered in both the Personal Records Database and the COVID-19-vaccination Information and Monitoring System (CIMS) on 18 September 2022 (the day before the vaccination campaign started) with at least one previous COVID-19 vaccination before the 2022 autumn booster campaign. The 2022 autumn booster campaign was the first autumn booster campaign targeted at persons aged ≥60 years and persons with underlying medical conditions, similar to subsequent years. For comparison, the analyses of the primary vaccination series were also performed among persons aged ≥60 years (results described in Supplement 6).

The study population was categorised into 14 foreign countries/regions of origin based on the classification of country of origin by Statistics Netherlands (CBS) [17]. Country/region of origin was defined based on an individual’s and their parents’ country of birth. The countries/regions for which the number of unvaccinated individuals was relatively high *and* vaccine uptake of the primary series was low were selected for the analyses (Supplement 1). The four thus selected countries/regions of origin were Morocco, Turkey, Suriname and Caribbean part of the Kingdom of the Netherlands. For comparison, the analyses were repeated for the population of Dutch origin.

### 2.2. Data sources

Vaccination data were obtained from CIMS. Vaccine uptake of the primary series was defined as having received at least one COVID-19 vaccination between the launch of the COVID-19 vaccination programme on 6 January 2021 and the start of the first booster vaccination campaign on 18 November 2021. Completion of the primary series could not be assessed, since previous infections are not registered in the CIMS database, while individuals with a prior SARS-CoV-2 infection required only one vaccination to complete the primary series in the Netherlands. Vaccine uptake of the 2022 autumn booster round was defined as having received a COVID-19 vaccination labelled as ‘booster’ between 19 September 2022 and 15 September 2023.

Vaccinated individuals who did not consent to registration of their vaccination in CIMS were included as unvaccinated. Individuals who did not consent to registration of their primary vaccination series (7% of vaccinees) were not included in the autumn booster analysis, as they were not eligible for booster vaccination [18]. In the 2022 autumn booster round, 99.2% consented to registration in CIMS (personal communication S. Lanooij).

Data on the potential determinants of being unvaccinated were obtained from CBS registers (Supplement 2), and selected based on previous studies [2, 11, 15, 19-21]. At individual level, we included: age, sex, migration background (whether a persons and/or their parents were born in the Netherlands), socioeconomic position (primary source of income) and medical risk group. Income, vehicle ownership, education level (highest completed education level of the household’s main earner or their partner), urbanisation level and distance to the nearest long-term vaccination location were included at household level. The determinant data were linked to the vaccination data at individual level within the remote access environment of the CBS.

### 2.3. Statistical analyses

To identify independent determinants and mediators, we built several multivariable logistic regression models by subsequently adding (sets of) variables to the previous model. The sets were based on the proximity of the determinants to the outcome, from most distal to most proximate. In order to avoid inappropriately adjusting odds ratios for mediating factors [22], we present the adjusted odds ratios (aORs) of non-uptake of vaccination of the model where the variable was first introduced. First, a model including age, sex and migration background was estimated. Socioeconomic position, household income and household education level were added in model 2, medical risk group in model 3, urbanisation level in model 4, household car ownership in model 5 and distance to the nearest long-term vaccination location in model 6. To assess whether the effect of migration background was explained by other variables, we examined the change in aOR of migration background as more variables were added to the model.

Furthermore, population attributable fractions (PAFs) were calculated. The PAFs estimate the percentage of non-uptake of COVID-19 vaccination that can be attributed to each determinant. The PAFs were calculated based on probabilities estimated through an adaptation of model 6, whereby some categories were merged and reference levels were adapted to ease interpretation of the PAFs. The PAFs were calculated as the change in probability of non-uptake when the value of the determinant was changed from the original level (P_0_) to the scenario level (P_scen_), i.e. the reference level:

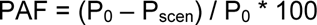

All analyses were conducted in RStudio, version 4.2.3.

### 2.4. Ethics statement

This study was deemed exempt from the law for medical research involving human subjects by the Centre for Clinical Expertise at the RIVM.

## 3. Results

### 3.1. Descriptive analyses

On 5 January 2021, the population of the Netherlands aged ≥18 years included 294480 persons of Moroccan origin, 333,090 of Turkish origin, 304,370 of Surinamese origin, 136,230 of Dutch-Caribbean origin and 10,660,430 of Dutch origin. The study population aged ≥60 years with at least one vaccination before the 2022 autumn booster round included 26,550 persons of Moroccan origin, 27,180 of Turkish origin, 54,750 of Surinamese origin, 16,900 of Dutch-Caribbean origin and 3,434,890 of Dutch origin. The 60-plus populations of Moroccan and Turkish origin for the analyses of the 2022 autumn booster round were limited to migrants with two foreign-born parents (Moroccan: 26,490 persons, Turkish: 27,060 persons), because the other migration background categories contained too few individuals.

Uptake of at least one COVID-19 vaccination in the primary vaccination round and in the 2022 autumn booster round varied between the countries of origin (uptake primary vaccination: Moroccan: 40.5%, Turkish: 53.4%, Surinamese: 62.9%, Dutch-Caribbean: 51.8%, Dutch: 84.9%, uptake autumn booster round: Moroccan: 8.3%, Turkish: 14.7%, Surinamese: 46.4%, Dutch-Caribbean: 50.6%, Dutch: 69.5%). Supplements 3 and 4 show vaccine uptake by determinant.

For several potential determinants of being unvaccinated, the distributions were similar in the populations of Turkish and Moroccan origin, but differed from the populations of Dutch-Caribbean and Surinamese origin (Table 1 & 2). In the study populations of both vaccination rounds, the proportion of people working in employment, income (only autumn booster round), education level, and vehicle ownership were lower in the former populations. In the study population of the primary vaccination round, over one-third of people of Moroccan (38.1%) and Turkish (36.8%) origin were born in the Netherlands to two foreign-born parents, compared to only 13.5% of people of Dutch-Caribbean origin. The distributions of sex, age, medical risk, urbanisation level and distance to vaccination location were broadly similar across the four populations of non-Dutch origin in both vaccination rounds.

**Table 1.**
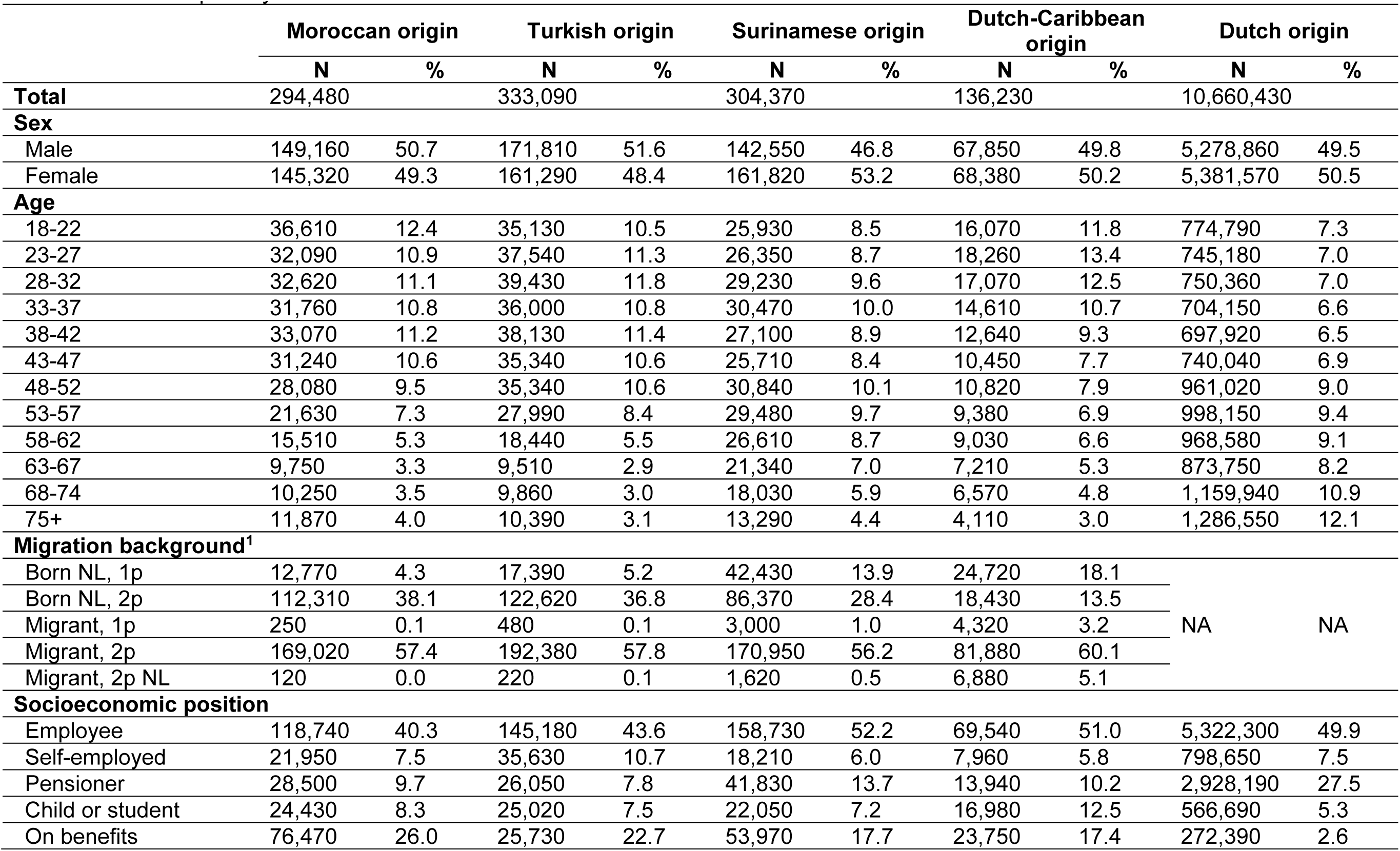
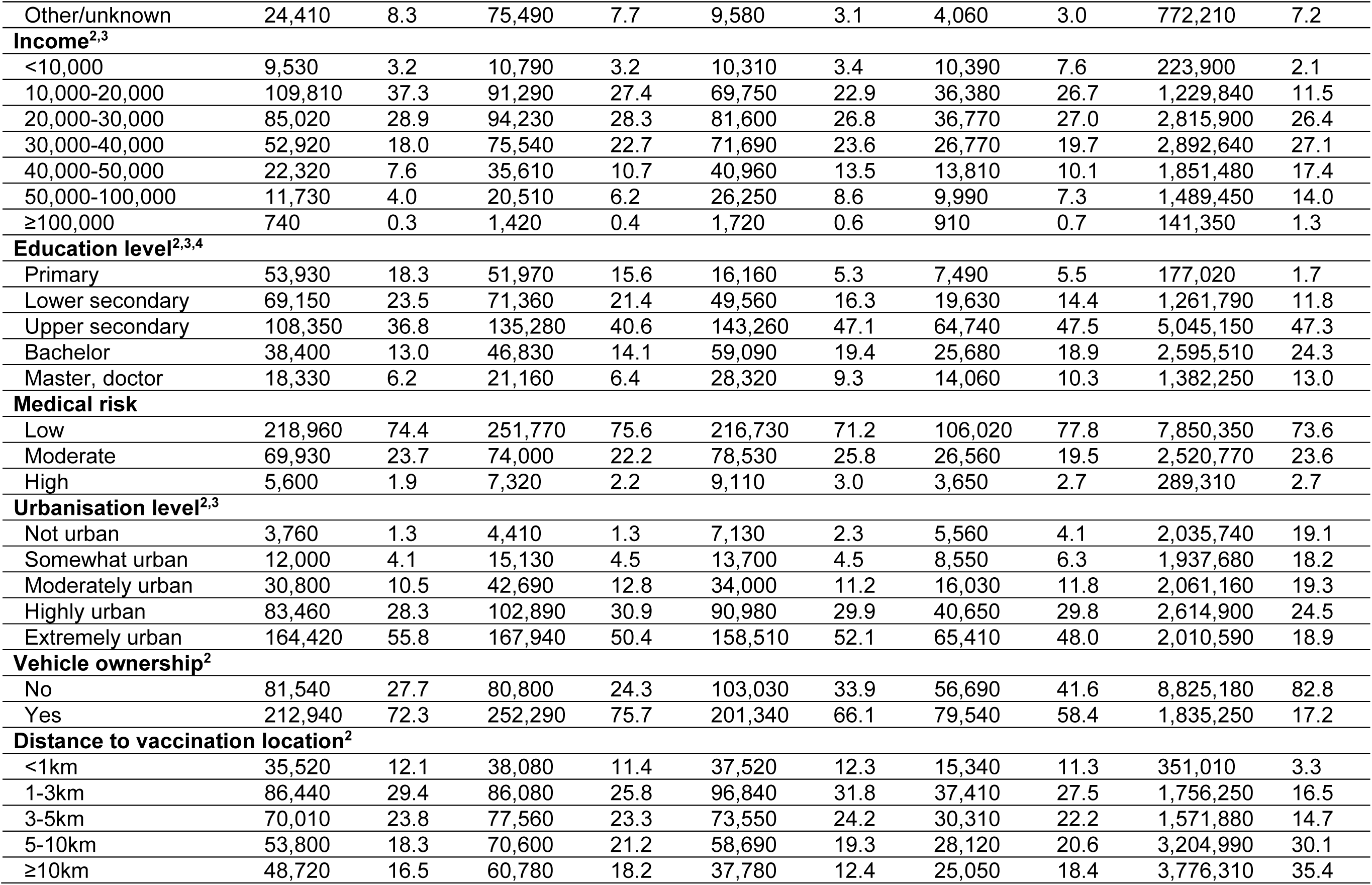
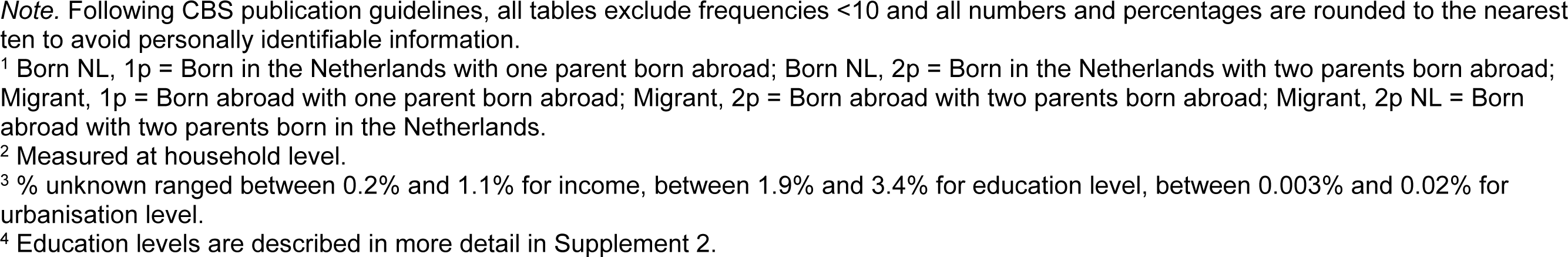
Characteristics of the populations by country of origin aged 18 years and older used for the analyses of determinants of being unvaccinated in the primary COVID-19 vaccination round.

|  | Moroccan origin |  | Turkish origin |  | Surinamese origin |  | Dutch-Caribbean origin |  | Dutch origin |  |
| --- | --- | --- | --- | --- | --- | --- | --- | --- | --- | --- |
|  | N | % | N | % | N | % | N | % | N | % |
| **Total** | 294,480 |  | 333,090 |  | 304,370 |  | 136,230 |  | 10,660,430 |  |
| **Sex** |  |  |  |  |  |  |  |  |  |  |
| Male | 149,160 | 50.7 | 171,810 | 51.6 | 142,550 | 46.8 | 67,850 | 49.8 | 5,278,860 | 49.5 |
| Female | 145,320 | 49.3 | 161,290 | 48.4 | 161,820 | 53.2 | 68,380 | 50.2 | 5,381,570 | 50.5 |
| **Age** |  |  |  |  |  |  |  |  |  |  |
| 18-22 | 36,610 | 12.4 | 35,130 | 10.5 | 25,930 | 8.5 | 16,070 | 11.8 | 774,790 | 7.3 |
| 23-27 | 32,090 | 10.9 | 37,540 | 11.3 | 26,350 | 8.7 | 18,260 | 13.4 | 745,180 | 7.0 |
| 28-32 | 32,620 | 11.1 | 39,430 | 11.8 | 29,230 | 9.6 | 17,070 | 12.5 | 750,360 | 7.0 |
| 33-37 | 31,760 | 10.8 | 36,000 | 10.8 | 30,470 | 10.0 | 14,610 | 10.7 | 704,150 | 6.6 |
| 38-42 | 33,070 | 11.2 | 38,130 | 11.4 | 27,100 | 8.9 | 12,640 | 9.3 | 697,920 | 6.5 |
| 43-47 | 31,240 | 10.6 | 35,340 | 10.6 | 25,710 | 8.4 | 10,450 | 7.7 | 740,040 | 6.9 |
| 48-52 | 28,080 | 9.5 | 35,340 | 10.6 | 30,840 | 10.1 | 10,820 | 7.9 | 961,020 | 9.0 |
| 53-57 | 21,630 | 7.3 | 27,990 | 8.4 | 29,480 | 9.7 | 9,380 | 6.9 | 998,150 | 9.4 |
| 58-62 | 15,510 | 5.3 | 18,440 | 5.5 | 26,610 | 8.7 | 9,030 | 6.6 | 968,580 | 9.1 |
| 63-67 | 9,750 | 3.3 | 9,510 | 2.9 | 21,340 | 7.0 | 7,210 | 5.3 | 873,750 | 8.2 |
| 68-74 | 10,250 | 3.5 | 9,860 | 3.0 | 18,030 | 5.9 | 6,570 | 4.8 | 1,159,940 | 10.9 |
| 75+ | 11,870 | 4.0 | 10,390 | 3.1 | 13,290 | 4.4 | 4,110 | 3.0 | 1,286,550 | 12.1 |
| **Migration background<sup>1</sup>** |  |  |  |  |  |  |  |  |  |  |
| Born NL, 1p | 12,770 | 4.3 | 17,390 | 5.2 | 42,430 | 13.9 | 24,720 | 18.1 | NA | NA |
| Born NL, 2p | 112,310 | 38.1 | 122,620 | 36.8 | 86,370 | 28.4 | 18,430 | 13.5 |  |  |
| Migrant, 1p | 250 | 0.1 | 480 | 0.1 | 3,000 | 1.0 | 4,320 | 3.2 |  |  |
| Migrant, 2p | 169,020 | 57.4 | 192,380 | 57.8 | 170,950 | 56.2 | 81,880 | 60.1 |  |  |
| Migrant, 2p NL | 120 | 0.0 | 220 | 0.1 | 1,620 | 0.5 | 6,880 | 5.1 |  |  |
| **Socioeconomic position** |  |  |  |  |  |  |  |  |  |  |
| Employee | 118,740 | 40.3 | 145,180 | 43.6 | 158,730 | 52.2 | 69,540 | 51.0 | 5,322,300 | 49.9 |
| Self-employed | 21,950 | 7.5 | 35,630 | 10.7 | 18,210 | 6.0 | 7,960 | 5.8 | 798,650 | 7.5 |
| Pensioner | 28,500 | 9.7 | 26,050 | 7.8 | 41,830 | 13.7 | 13,940 | 10.2 | 2,928,190 | 27.5 |
| Child or student | 24,430 | 8.3 | 25,020 | 7.5 | 22,050 | 7.2 | 16,980 | 12.5 | 566,690 | 5.3 |
| On benefits | 76,470 | 26.0 | 25,730 | 22.7 | 53,970 | 17.7 | 23,750 | 17.4 | 272,390 | 2.6 |
| Other/unknown | 24,410 | 8.3 | 75,490 | 7.7 | 9,580 | 3.1 | 4,060 | 3.0 | 772,210 | 7.2 |
| **Income<sup>2,3</sup>** |  |  |  |  |  |  |  |  |  |  |
| <10,000 | 9,530 | 3.2 | 10,790 | 3.2 | 10,310 | 3.4 | 10,390 | 7.6 | 223,900 | 2.1 |
| 10,000-20,000 | 109,810 | 37.3 | 91,290 | 27.4 | 69,750 | 22.9 | 36,380 | 26.7 | 1,229,840 | 11.5 |
| 20,000-30,000 | 85,020 | 28.9 | 94,230 | 28.3 | 81,600 | 26.8 | 36,770 | 27.0 | 2,815,900 | 26.4 |
| 30,000-40,000 | 52,920 | 18.0 | 75,540 | 22.7 | 71,690 | 23.6 | 26,770 | 19.7 | 2,892,640 | 27.1 |
| 40,000-50,000 | 22,320 | 7.6 | 35,610 | 10.7 | 40,960 | 13.5 | 13,810 | 10.1 | 1,851,480 | 17.4 |
| 50,000-100,000 | 11,730 | 4.0 | 20,510 | 6.2 | 26,250 | 8.6 | 9,990 | 7.3 | 1,489,450 | 14.0 |
| ≥100,000 | 740 | 0.3 | 1,420 | 0.4 | 1,720 | 0.6 | 910 | 0.7 | 141,350 | 1.3 |
| **Education level<sup>2,3,4</sup>** |  |  |  |  |  |  |  |  |  |  |
| Primary | 53,930 | 18.3 | 51,970 | 15.6 | 16,160 | 5.3 | 7,490 | 5.5 | 177,020 | 1.7 |
| Lower secondary | 69,150 | 23.5 | 71,360 | 21.4 | 49,560 | 16.3 | 19,630 | 14.4 | 1,261,790 | 11.8 |
| Upper secondary | 108,350 | 36.8 | 135,280 | 40.6 | 143,260 | 47.1 | 64,740 | 47.5 | 5,045,150 | 47.3 |
| Bachelor | 38,400 | 13.0 | 46,830 | 14.1 | 59,090 | 19.4 | 25,680 | 18.9 | 2,595,510 | 24.3 |
| Master, doctor | 18,330 | 6.2 | 21,160 | 6.4 | 28,320 | 9.3 | 14,060 | 10.3 | 1,382,250 | 13.0 |
| **Medical risk** |  |  |  |  |  |  |  |  |  |  |
| Low | 218,960 | 74.4 | 251,770 | 75.6 | 216,730 | 71.2 | 106,020 | 77.8 | 7,850,350 | 73.6 |
| Moderate | 69,930 | 23.7 | 74,000 | 22.2 | 78,530 | 25.8 | 26,560 | 19.5 | 2,520,770 | 23.6 |
| High | 5,600 | 1.9 | 7,320 | 2.2 | 9,110 | 3.0 | 3,650 | 2.7 | 289,310 | 2.7 |
| **Urbanisation level<sup>2,3</sup>** |  |  |  |  |  |  |  |  |  |  |
| Not urban | 3,760 | 1.3 | 4,410 | 1.3 | 7,130 | 2.3 | 5,560 | 4.1 | 2,035,740 | 19.1 |
| Somewhat urban | 12,000 | 4.1 | 15,130 | 4.5 | 13,700 | 4.5 | 8,550 | 6.3 | 1,937,680 | 18.2 |
| Moderately urban | 30,800 | 10.5 | 42,690 | 12.8 | 34,000 | 11.2 | 16,030 | 11.8 | 2,061,160 | 19.3 |
| Highly urban | 83,460 | 28.3 | 102,890 | 30.9 | 90,980 | 29.9 | 40,650 | 29.8 | 2,614,900 | 24.5 |
| Extremely urban | 164,420 | 55.8 | 167,940 | 50.4 | 158,510 | 52.1 | 65,410 | 48.0 | 2,010,590 | 18.9 |
| **Vehicle ownership<sup>2</sup>** |  |  |  |  |  |  |  |  |  |  |
| No | 81,540 | 27.7 | 80,800 | 24.3 | 103,030 | 33.9 | 56,690 | 41.6 | 8,825,180 | 82.8 |
| Yes | 212,940 | 72.3 | 252,290 | 75.7 | 201,340 | 66.1 | 79,540 | 58.4 | 1,835,250 | 17.2 |
| **Distance to vaccination location<sup>2</sup>** |  |  |  |  |  |  |  |  |  |  |
| <1km | 35,520 | 12.1 | 38,080 | 11.4 | 37,520 | 12.3 | 15,340 | 11.3 | 351,010 | 3.3 |
| 1-3km | 86,440 | 29.4 | 86,080 | 25.8 | 96,840 | 31.8 | 37,410 | 27.5 | 1,756,250 | 16.5 |
| 3-5km | 70,010 | 23.8 | 77,560 | 23.3 | 73,550 | 24.2 | 30,310 | 22.2 | 1,571,880 | 14.7 |
| 5-10km | 53,800 | 18.3 | 70,600 | 21.2 | 58,690 | 19.3 | 28,120 | 20.6 | 3,204,990 | 30.1 |
| ≥10km | 48,720 | 16.5 | 60,780 | 18.2 | 37,780 | 12.4 | 25,050 | 18.4 | 3,776,310 | 35.4 |

*Note.* Following CBS publication guidelines, all tables exclude frequencies <10 and all numbers and percentages are rounded to the nearest ten to avoid personally identifiable information.
<sup>1</sup> Born NL, 1p = Born in the Netherlands with one parent born abroad; Born NL, 2p = Born in the Netherlands with two parents born abroad; Migrant, 1p = Born abroad with one parent born abroad; Migrant, 2p = Born abroad with two parents born abroad; Migrant, 2p NL = Born abroad with two parents born in the Netherlands.
<sup>2</sup> Measured at household level.
<sup>3</sup> % unknown ranged between 0.2% and 1.1% for income, between 1.9% and 3.4% for education level, between 0.003% and 0.02% for urbanisation level.
<sup>4</sup> Education levels are described in more detail in Supplement 2.

**Table 2.**
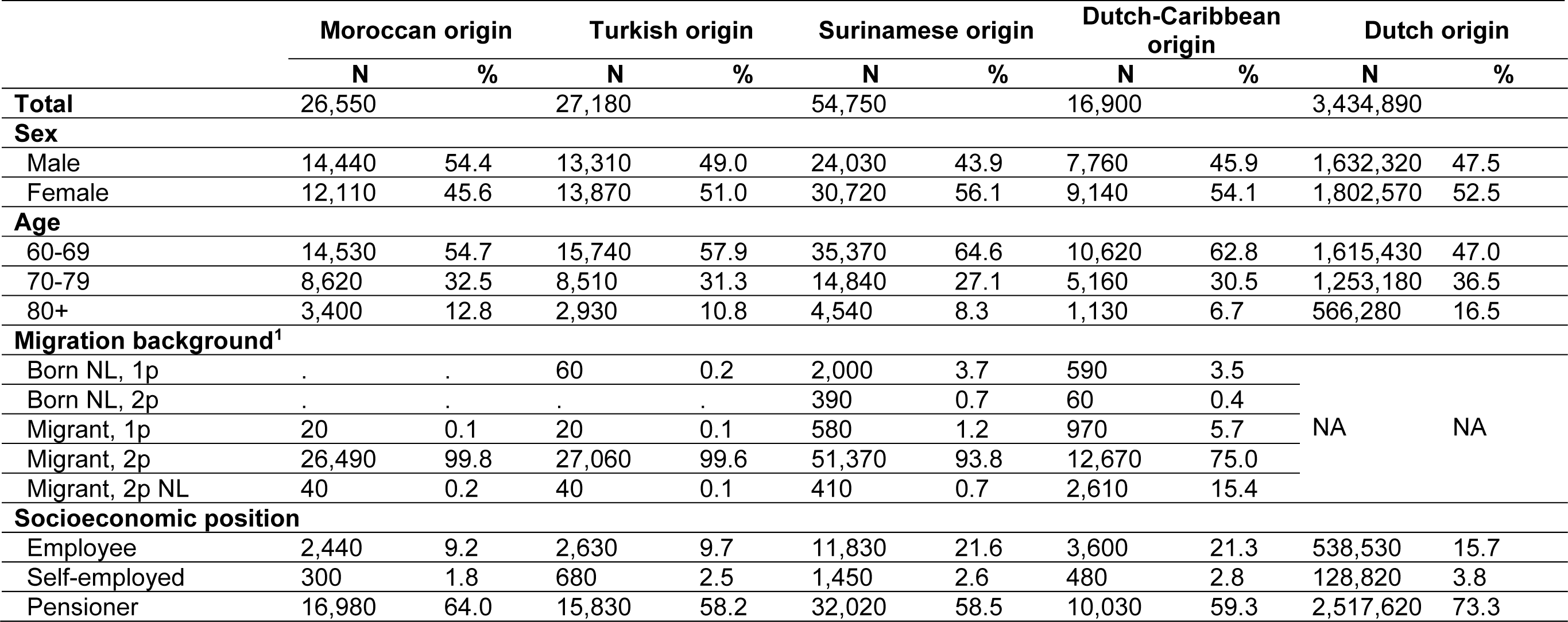
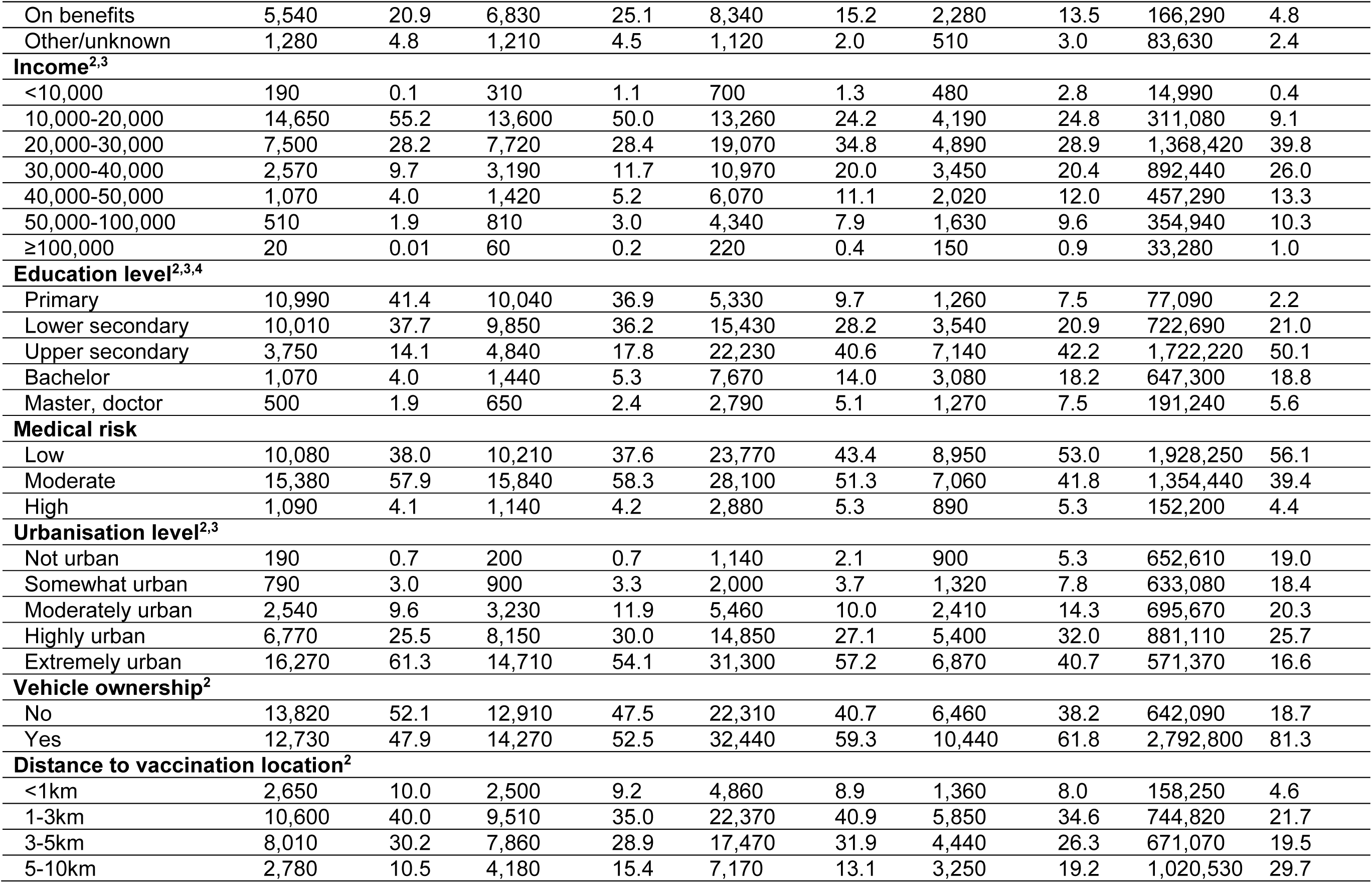
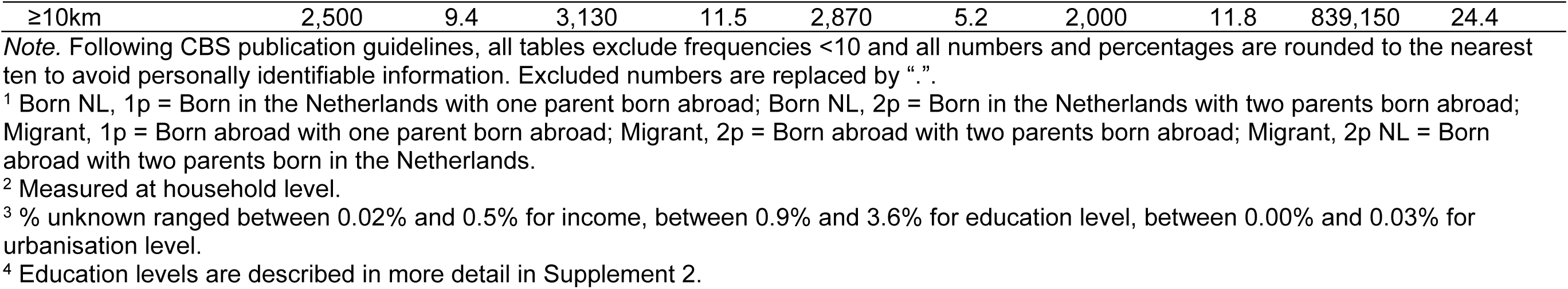
Characteristics of the populations by country of origin aged 60 years and older used for the analyses of determinants of being unvaccinated in the autumn 2022 COVID-19 booster round.

|  | Moroccan origin |  | Turkish origin |  | Surinamese origin |  | Dutch-Caribbean origin |  | Dutch origin |  |
| --- | --- | --- | --- | --- | --- | --- | --- | --- | --- | --- |
|  | N | % | N | % | N | % | N | % | N | % |
| **Total** | 26,550 |  | 27,180 |  | 54,750 |  | 16,900 |  | 3,434,890 |  |
| **Sex** |  |  |  |  |  |  |  |  |  |  |
| Male | 14,440 | 54.4 | 13,310 | 49.0 | 24,030 | 43.9 | 7,760 | 45.9 | 1,632,320 | 47.5 |
| Female | 12,110 | 45.6 | 13,870 | 51.0 | 30,720 | 56.1 | 9,140 | 54.1 | 1,802,570 | 52.5 |
| **Age** |  |  |  |  |  |  |  |  |  |  |
| 60-69 | 14,530 | 54.7 | 15,740 | 57.9 | 35,370 | 64.6 | 10,620 | 62.8 | 1,615,430 | 47.0 |
| 70-79 | 8,620 | 32.5 | 8,510 | 31.3 | 14,840 | 27.1 | 5,160 | 30.5 | 1,253,180 | 36.5 |
| 80+ | 3,400 | 12.8 | 2,930 | 10.8 | 4,540 | 8.3 | 1,130 | 6.7 | 566,280 | 16.5 |
| **Migration background<sup>1</sup>** |  |  |  |  |  |  |  |  |  |  |
| Born NL, 1p | . | . | 60 | 0.2 | 2,000 | 3.7 | 590 | 3.5 | NA | NA |
| Born NL, 2p | . | . | . | . | 390 | 0.7 | 60 | 0.4 |  |  |
| Migrant, 1p | 20 | 0.1 | 20 | 0.1 | 580 | 1.2 | 970 | 5.7 |  |  |
| Migrant, 2p | 26,490 | 99.8 | 27,060 | 99.6 | 51,370 | 93.8 | 12,670 | 75.0 |  |  |
| Migrant, 2p NL | 40 | 0.2 | 40 | 0.1 | 410 | 0.7 | 2,610 | 15.4 |  |  |
| **Socioeconomic position** |  |  |  |  |  |  |  |  |  |  |
| Employee | 2,440 | 9.2 | 2,630 | 9.7 | 11,830 | 21.6 | 3,600 | 21.3 | 538,530 | 15.7 |
| Self-employed | 300 | 1.8 | 680 | 2.5 | 1,450 | 2.6 | 480 | 2.8 | 128,820 | 3.8 |
| Pensioner | 16,980 | 64.0 | 15,830 | 58.2 | 32,020 | 58.5 | 10,030 | 59.3 | 2,517,620 | 73.3 |
| On benefits | 5,540 | 20.9 | 6,830 | 25.1 | 8,340 | 15.2 | 2,280 | 13.5 | 166,290 | 4.8 |
| Other/unknown | 1,280 | 4.8 | 1,210 | 4.5 | 1,120 | 2.0 | 510 | 3.0 | 83,630 | 2.4 |
| **Income<sup>2,3</sup>** |  |  |  |  |  |  |  |  |  |  |
| <10,000 | 190 | 0.1 | 310 | 1.1 | 700 | 1.3 | 480 | 2.8 | 14,990 | 0.4 |
| 10,000-20,000 | 14,650 | 55.2 | 13,600 | 50.0 | 13,260 | 24.2 | 4,190 | 24.8 | 311,080 | 9.1 |
| 20,000-30,000 | 7,500 | 28.2 | 7,720 | 28.4 | 19,070 | 34.8 | 4,890 | 28.9 | 1,368,420 | 39.8 |
| 30,000-40,000 | 2,570 | 9.7 | 3,190 | 11.7 | 10,970 | 20.0 | 3,450 | 20.4 | 892,440 | 26.0 |
| 40,000-50,000 | 1,070 | 4.0 | 1,420 | 5.2 | 6,070 | 11.1 | 2,020 | 12.0 | 457,290 | 13.3 |
| 50,000-100,000 | 510 | 1.9 | 810 | 3.0 | 4,340 | 7.9 | 1,630 | 9.6 | 354,940 | 10.3 |
| ≥100,000 | 20 | 0.01 | 60 | 0.2 | 220 | 0.4 | 150 | 0.9 | 33,280 | 1.0 |
| **Education level<sup>2,3,4</sup>** |  |  |  |  |  |  |  |  |  |  |
| Primary | 10,990 | 41.4 | 10,040 | 36.9 | 5,330 | 9.7 | 1,260 | 7.5 | 77,090 | 2.2 |
| Lower secondary | 10,010 | 37.7 | 9,850 | 36.2 | 15,430 | 28.2 | 3,540 | 20.9 | 722,690 | 21.0 |
| Upper secondary | 3,750 | 14.1 | 4,840 | 17.8 | 22,230 | 40.6 | 7,140 | 42.2 | 1,722,220 | 50.1 |
| Bachelor | 1,070 | 4.0 | 1,440 | 5.3 | 7,670 | 14.0 | 3,080 | 18.2 | 647,300 | 18.8 |
| Master, doctor | 500 | 1.9 | 650 | 2.4 | 2,790 | 5.1 | 1,270 | 7.5 | 191,240 | 5.6 |
| **Medical risk** |  |  |  |  |  |  |  |  |  |  |
| Low | 10,080 | 38.0 | 10,210 | 37.6 | 23,770 | 43.4 | 8,950 | 53.0 | 1,928,250 | 56.1 |
| Moderate | 15,380 | 57.9 | 15,840 | 58.3 | 28,100 | 51.3 | 7,060 | 41.8 | 1,354,440 | 39.4 |
| High | 1,090 | 4.1 | 1,140 | 4.2 | 2,880 | 5.3 | 890 | 5.3 | 152,200 | 4.4 |
| **Urbanisation level<sup>2,3</sup>** |  |  |  |  |  |  |  |  |  |  |
| Not urban | 190 | 0.7 | 200 | 0.7 | 1,140 | 2.1 | 900 | 5.3 | 652,610 | 19.0 |
| Somewhat urban | 790 | 3.0 | 900 | 3.3 | 2,000 | 3.7 | 1,320 | 7.8 | 633,080 | 18.4 |
| Moderately urban | 2,540 | 9.6 | 3,230 | 11.9 | 5,460 | 10.0 | 2,410 | 14.3 | 695,670 | 20.3 |
| Highly urban | 6,770 | 25.5 | 8,150 | 30.0 | 14,850 | 27.1 | 5,400 | 32.0 | 881,110 | 25.7 |
| Extremely urban | 16,270 | 61.3 | 14,710 | 54.1 | 31,300 | 57.2 | 6,870 | 40.7 | 571,370 | 16.6 |
| **Vehicle ownership<sup>2</sup>** |  |  |  |  |  |  |  |  |  |  |
| No | 13,820 | 52.1 | 12,910 | 47.5 | 22,310 | 40.7 | 6,460 | 38.2 | 642,090 | 18.7 |
| Yes | 12,730 | 47.9 | 14,270 | 52.5 | 32,440 | 59.3 | 10,440 | 61.8 | 2,792,800 | 81.3 |
| **Distance to vaccination location<sup>2</sup>** |  |  |  |  |  |  |  |  |  |  |
| <1km | 2,650 | 10.0 | 2,500 | 9.2 | 4,860 | 8.9 | 1,360 | 8.0 | 158,250 | 4.6 |
| 1-3km | 10,600 | 40.0 | 9,510 | 35.0 | 22,370 | 40.9 | 5,850 | 34.6 | 744,820 | 21.7 |
| 3-5km | 8,010 | 30.2 | 7,860 | 28.9 | 17,470 | 31.9 | 4,440 | 26.3 | 671,070 | 19.5 |
| 5-10km | 2,780 | 10.5 | 4,180 | 15.4 | 7,170 | 13.1 | 3,250 | 19.2 | 1,020,530 | 29.7 |
| ≥10km | 2,500 | 9.4 | 3,130 | 11.5 | 2,870 | 5.2 | 2,000 | 11.8 | 839,150 | 24.4 |

*Note.* Following CBS publication guidelines, all tables exclude frequencies <10 and all numbers and percentages are rounded to the nearest ten to avoid personally identifiable information. Excluded numbers are replaced by “.”.
<sup>1</sup> Born NL, 1p = Born in the Netherlands with one parent born abroad; Born NL, 2p = Born in the Netherlands with two parents born abroad; Migrant, 1p = Born abroad with one parent born abroad; Migrant, 2p = Born abroad with two parents born abroad; Migrant, 2p NL = Born abroad with two parents born in the Netherlands.
<sup>2</sup> Measured at household level.
<sup>3</sup> % unknown ranged between 0.02% and 0.5% for income, between 0.9% and 3.6% for education level, between 0.00% and 0.03% for urbanisation level.
<sup>4</sup> Education levels are described in more detail in Supplement 2.

### 3.2. Determinants of being unvaccinated in the primary vaccination round

For many determinants, the estimated effects were similar in all four populations of non-Dutch origin, but some differences were observed (Table 3). Overall, the odds of being unvaccinated in the primary vaccination round among persons aged ≥18 years increased as age decreased. This trend was observed for age groups below 63, 58 and 48 years in the populations of Surinamese, Moroccan and Turkish origin, respectively. The odds of being unvaccinated were particularly high among young persons of Dutch-Caribbean or Moroccan origin (aORs >5 below 38 and 33 years, respectively). The odds of being unvaccinated in the primary vaccination round increased as income and education level decreased. The effect of education was strongest in the population of Dutch-Caribbean origin. Being self-employed (aORs between 1.53 and 1.75), a pensioner (aORs between 1.09 and 1.80), on benefits (aORs between 1.34 and 1.47) or having other/unknown socioeconomic position (aORs between 1.51 and 1.86) were associated with lower uptake in nearly all populations of non-Dutch origin. Moderate or no medical risk compared to high medical risk was also associated with lower uptake. Similar effects of age, income, education level, socioeconomic position and medical risk were observed among persons of Dutch origin.

**Table 3.** Results of multilevel multivariable models of the association between determinants and being unvaccinated in the primary COVID-19 vaccination round among persons aged 18 years and older of Moroccan, Turkish, Surinamese, Dutch-Caribbean and Dutch origin.

|  | Moroccan origin |  | Turkish origin |  | Surinamese origin |  | Dutch-Caribbean origin |  | Dutch origin |  |
| --- | --- | --- | --- | --- | --- | --- | --- | --- | --- | --- |
|  | aOR | 95% CI | aOR | 95% CI | aOR | 95% CI | aOR | 95% CI | aOR | 95% CI |
| **Sex<sup>1</sup>** |  |  |  |  |  |  |  |  |  |  |
| Male | (ref) |  | (ref) |  | (ref) |  | (ref) |  | (ref) |  |
| Female | **1.10** | 1.08-1.12 | 1.00 | 0.99-1.02 | **0.97** | 0.96-0.99 | 1.01 | 0.99-1.03 | **0.95** | 0.95-0.96 |
| **Age<sup>2</sup>** |  |  |  |  |  |  |  |  |  |  |
| 75+ | (ref) |  | (ref) |  | (ref) |  | (ref) |  | (ref) |  |
| 68-74 | **0.87** | 0.82-0.92 | **1.10** | 1.04-1.17 | **0.91** | 0.86-0.97 | **1.16** | 1.06-1.28 | **0.76** | 0.76-0.77 |
| 63-67 | **0.85** | 0.80-0.90 | 0.99 | 0.94-1.06 | **0.94** | 0.89-1.00 | **1.35** | 1.23-1.47 | **0.88** | 0.88-0.89 |
| 58-62 | 0.95 | 0.91-1.00 | **0.87** | 0.83-0.92 | **1.07** | 1.02-1.13 | **1.61** | 1.47-1.75 | **1.07** | 1.06-1.07 |
| 53-57 | **1.09** | 1.04-1.14 | **0.91** | 0.87-0.96 | **1.37** | 1.30-1.44 | **2.03** | 1.87-2.21 | **1.37** | 1.36-1.38 |
| 48-52 | **1.32** | 1.26-1.38 | 0.98 | 0.94-1.03 | **1.63** | 1.55-1.71 | **2.45** | 2.25-2.67 | **1.61** | 1.60-1.63 |
| 43-47 | **1.59** | 1.52-1.66 | **1.09** | 1.04-1.14 | **1.96** | 1.86-2.06 | **2.97** | 2.73-3.24 | **1.87** | 1.85-1.89 |
| 38-42 | **2.14** | 2.05-2.24 | **1.24** | 1.18-1.30 | **2.70** | 2.56-2.84 | **3.78** | 3.48-4.11 | **2.33** | 2.31-2.35 |
| 33-37 | **3.37** | 3.22-3.53 | **1.81** | 1.73-1.90 | **3.69** | 3.51-3.88 | **5.24** | 4.83-5.69 | **2.97** | 2.95-3.00 |
| 28-32 | **5.33** | 5.08-5.60 | **2.65** | 2.52-2.78 | **4.73** | 4.50-4.98 | **6.40** | 5.90-6.95 | **3.54** | 3.51-3.57 |
| 23-27 | **6.51** | 6.18-6.85 | **3.57** | 3.39-3.75 | **5.00** | 4.74-5.26 | **6.62** | 6.10-7.18 | **3.39** | 3.36-3.42 |
| 18-22 | **7.06** | 6.70-7.43 | **3.83** | 3.64-4.03 | **4.09** | 3.88-4.32 | **6.71** | 6.18-7.29 | **2.91** | 2.88-2.93 |
| **Migration background<sup>3</sup>** |  |  |  |  |  |  |  |  |  |  |
| Born NL, 1p | (ref) |  | (ref) |  | (ref) |  | (ref) |  |  |  |
| Born NL, 2p | **2.71** | 2.60-2.81 | **1.58** | 1.52-1.63 | **1.56** | 1.52-1.60 | **3.67** | 3.52-3.83 |  |  |
| Migrant, 1p | 1.22 | 0.93-1.59 | **0.73** | 0.61-0.88 | **1.17** | 1.08-1.27 | 0.98 | 0.91-1.06 | NA | NA |
| Migrant, 2p | **2.16** | 2.07-2.26 | **1.10** | 1.06-1.14 | **1.21** | 1.18-1.25 | **3.39** | 3.28-3.50 |  |  |
| Migrant, 2p NL | 0.70 | 0.46-1.07 | **0.61** | 0.45-0.81 | **0.62** | 0.55-0.71 | **0.56** | 0.52-0.60 |  |  |
| **Socioeconomic position<sup>4</sup>** |  |  |  |  |  |  |  |  |  |  |
| Employee | (ref) |  | (ref) |  | (ref) |  | (ref) |  | (ref) |  |
| Self-employed | **1.61** | 1.56-1.67 | **1.75** | 1.71-1.80 | **1.53** | 1.48-1.58 | **1.73** | 1.64-1.82 | **1.96** | 1.95-1.97 |
| Pensioner | **1.59** | 1.50-1.69 | **1.80** | 1.69-1.91 | **1.18** | 1.11-1.25 | 1.09 | 0.99-1.19 | **1.18** | 1.16-1.19 |
| Child or student | **0.77** | 0.74-0.81 | **0.74** | 0.72-0.77 | **0.63** | 0.60-0.65 | **0.69** | 0.65-0.73 | **0.48** | 0.48-0.49 |
| On benefits | **1.45** | 1.41-1.48 | **1.47** | 1.44-1.50 | **1.34** | 1.31-1.37 | **1.44** | 1.39-1.50 | **1.66** | 1.65-1.68 |
| Other/unknown | **1.57** | 1.52-1.63 | **1.86** | 1.80-1.92 | **1.51** | 1.43-1.59 | **1.72** | 1.58-1.88 | **1.95** | 1.93-1.97 |
| **Income<sup>5</sup>** |  |  |  |  |  |  |  |  |  |  |
| ≥100,000 | (ref) |  | (ref) |  | (ref) |  | (ref) |  | (ref) |  |
| 50,000-100,000 | 1.02 | 0.87-1.20 | 1.05 | 0.93-1.18 | **1.31** | 1.15-1.50 | **1.23** | 1.01-1.52 | 0.99 | 0.97-1.01 |
| 40,000-50,000 | **1.22** | 1.04-1.44 | **1.14** | 1.01-1.29 | **1.53** | 1.35-1.75 | **1.67** | 1.37-2.06 | **1.08** | 1.06-1.10 |
| 30,000-40,000 | **1.34** | 1.14-1.57 | **1.26** | 1.12-1.43 | **1.80** | 1.59-2.06 | **1.94** | 1.59-2.38 | **1.23** | 1.21-1.26 |
| 20,000-30,000 | **1.50** | 1.28-1.76 | **1.39** | 1.24-1.57 | **2.35** | 2.06-2.67 | **2.49** | 2.04-3.06 | **1.56** | 1.53-1.59 |
| 10,000-20,000 | **1.58** | 1.35-1.85 | **1.53** | 1.36-1.72 | **2.91** | 2.56-3.33 | **2.98** | 2.44-3.67 | **2.19** | 2.15-2.24 |
| <10,000 | **2.01** | 1.70-2.37 | **1.99** | 1.76-2.26 | **3.07** | 2.68-3.52 | **2.36** | 1.93-2.92 | **2.15** | 2.11-2.20 |
| **Education level<sup>6</sup>** |  |  |  |  |  |  |  |  |  |  |
| Master, doctor | (ref) |  | (ref) |  | (ref) |  | (ref) |  | (ref) |  |
| Bachelor | **1.38** | 1.33-1.43 | **1.40** | 1.35-1.45 | **1.46** | 1.41-1.51 | **1.67** | 1.59-1.75 | **1.51** | 1.50-1.52 |
| Upper secondary | **1.58** | 1.53-1.64 | **1.58** | 1.53-1.63 | **1.74** | 1.68-1.79 | **2.46** | 2.34-2.58 | **2.03** | 2.02-2.05 |
| Lower secondary | **1.65** | 1.59-1.71 | **1.60** | 1.54-1.65 | **1.88** | 1.81-1.95 | **2.97** | 2.81-3.15 | **2.58** | 2.55-2.60 |
| Primary | **1.82** | 1.75-1.90 | **1.79** | 1.72-1.85 | **1.74** | 1.66-1.83 | **3.41** | 3.18-3.67 | **3.14** | 3.10-3.18 |
| **Medical risk<sup>7</sup>** |  |  |  |  |  |  |  |  |  |  |
| High risk | (ref) |  | (ref) |  | (ref) |  | (ref) |  | (ref) |  |
| Moderate risk | **1.36** | 1.28-1.44 | **1.38** | 1.31-1.46 | **1.12** | 1.06-1.18 | **1.35** | 1.24-1.46 | **1.08** | 1.06-1.09 |
| None | **1.75** | 1.65-1.85 | **1.74** | 1.65-1.84 | **1.71** | 1.63-1.80 | **1.67** | 1.55-1.81 | **1.35** | 1.33-1.36 |
| **Urbanisation level<sup>8</sup>** |  |  |  |  |  |  |  |  |  |  |
| Not urban | (ref) |  | (ref) |  | (ref) |  | (ref) |  | (ref) |  |
| Somewhat urban | **1.21** | 1.11-1.32 | 1.01 | 0.94-1.09 | 0.98 | 0.91-1.04 | **0.92** | 0.84-1.00 | **0.86** | 0.86-0.87 |
| Moderately urban | **1.20** | 1.11-1.30 | 1.05 | 0.98-1.13 | 1.00 | 0.94-1.06 | 0.96 | 0.89-1.04 | **0.86** | 0.85-0.86 |
| Highly urban | **1.31** | 1.21-1.41 | **1.14** | 1.06-1.23 | **1.16** | 1.10-1.23 | **1.10** | 1.02-1.17 | **0.92** | 0.91-0.92 |
| Extremely urban | **1.47** | 1.36-1.59 | **1.29** | 1.20-1.39 | **1.15** | 1.09-1.22 | 1.07 | 0.99-1.14 | **0.82** | 0.81-0.82 |
| **Vehicle ownership<sup>9</sup>** |  |  |  |  |  |  |  |  |  |  |
| No | (ref) |  | (ref) |  | (ref) |  | (ref) |  | (ref) |  |
| Yes | **1.08** | 1.06-1.10 | **1.09** | 1.07-1.11 | **0.82** | 0.80-0.84 | **1.13** | 1.09-1.16 | **0.99** | 0.98-1.00 |
| **Distance to vaccination location<sup>10</sup>** |  |  |  |  |  |  |  |  |  |  |
| <1km | (ref) |  | (ref) |  | (ref) |  | (ref) |  | (ref) |  |
| 1-3km | **0.92** | 0.90-0.95 | **0.84** | 0.81-0.86 | **0.92** | 0.90-0.95 | **0.85** | 0.81-0.89 | **0.92** | 0.91-0.93 |
| 3-5km | **0.91** | 0.89-0.94 | **0.83** | 0.81-0.85 | **0.81** | 0.79-0.84 | **0.82** | 0.79-0.86 | **0.96** | 0.95-0.97 |
| 5-10km | **0.92** | 0.89-0.95 | **0.88** | 0.85-0.90 | **0.83** | 0.80-0.85 | **0.84** | 0.80-0.88 | **0.97** | 0.96-0.98 |
| ≥10km | **0.86** | 0.83-0.89 | **0.81** | 0.79-0.84 | **0.82** | 0.79-0.85 | **0.82** | 0.78-0.86 | 1.00 | 0.99-1.01 |

<sup>1</sup> Adjusted for age and migration background (populations of Dutch-Caribbean and Surinamese origin only).
<sup>2</sup> Adjusted for sex and migration background (populations of Dutch-Caribbean and Surinamese origin only).
<sup>3</sup> Adjusted for age and sex. Born NL, 1p = Born in the Netherlands with one parent born abroad; Born NL, 2p = Born in the Netherlands with two parents born abroad; Migrant, 1p = Born abroad with one parent born abroad; Migrant, 2p = Born abroad with two parents born abroad; Migrant, 2p NL = Born abroad with two parents born in the Netherlands. For the populations of Moroccan, Surinamese and Turkish origin, the migrant categories were merged into: Migrant = Born abroad. Not applicable (N.A.) for the population of Dutch origin.
<sup>4</sup> Adjusted for age, sex, migration background (populations of Dutch-Caribbean and Surinamese origin only), income and education level.
<sup>5</sup> Measured at household level. Adjusted for age, sex, migration background (populations of Dutch-Caribbean and Surinamese origin only), socioeconomic position and education level.
<sup>6</sup> Measured at household level. Adjusted for age, sex, migration background (populations of Dutch-Caribbean and Surinamese origin only), socioeconomic position and income. Education levels are described in more detail in Supplement 2.
<sup>7</sup> Adjusted for age, sex, migration background (populations of Dutch-Caribbean and Surinamese origin only), socioeconomic position, income and education level.
<sup>8</sup> Measured at household level. Adjusted for age, sex, migration background (populations of Dutch-Caribbean and Surinamese origin only), medical risk, socioeconomic position, income and education level.
<sup>9</sup> Measured at household level. Adjusted for age, sex, migration background (populations of Dutch-Caribbean and Surinamese origin only), medical risk, socioeconomic position, income, education level and urbanisation level.
<sup>10</sup> Measured at household level. Adjusted for age, sex, migration background (populations of Dutch-Caribbean and Surinamese origin only), medical risk, socioeconomic position, income, education level, urbanisation level and vehicle ownership.

Being a migrant (i.e. born abroad) or being born in the Netherlands with two foreign-born parents, compared to being born in the Netherlands with one foreign-born parent, increased the odds of being unvaccinated in the primary vaccination round in all four populations of non-Dutch origin (born in the Netherlands with two foreign-born parents: aORs between 1.56 and 3.67, migrant with two foreign-born parents: aORs between 1.10 and 3.39). The effect of being a migrant with two parents born abroad on the odds of being unvaccinated was reduced by adding socioeconomic position, income and education level to the model but hardly changed after adding any of the other variables in the populations of Dutch-Caribbean and Moroccan origin (Supplement 5 Table S5.1). This indicates that the effect of being a migrant with two foreign-born parents is partially mediated by socioeconomic status. Among people of Turkish origin, the effect was weak (aOR=1.10) and inverted after adjustment for socioeconomic variables. For persons of Surinamese origin, the rather weak association (aOR=1.21) weakened further after adjustment for socioeconomic variables. The association between being a migrant with one foreign-born parent and being unvaccinated in the primary vaccination round varied between populations from different countries of origin.

The associations between vaccine uptake and sex, urbanisation level, vehicle ownership and distance to vaccination location were weaker or more ambiguous. Living in a highly or extremely urban area compared to a rural area were generally associated with lower uptake (aORs between 1.07 and 1.47) in the populations of non-Dutch origin, while the effect of living in a somewhat or moderately urban area varied between countries of origin. The effect of urbanisation was strongest among persons of Moroccan origin, for whom the odds of being unvaccinated increased with each urbanisation level (aORs from 1.21 to 1.47). This is in contrast to the population of Dutch origin, whereby higher urbanisation was associated with higher uptake. Vehicle ownership was weakly associated with lower uptake in all groups of non-Dutch origin (aORs between 1.08 and 1.13), except in the population of Surinamese origin (aOR=0.82). Among persons of Dutch origin the effect of vehicle ownership on being unvaccinated was negligible (aOR=0.99). The associations between vaccine uptake and distance to vaccination location were weak and no clear patterns were observed.

### 3.3 Determinants of being unvaccinated in the autumn 2022 booster round for individuals aged 60 years and older

Due to smaller study populations of people aged ≥60 years and the distribution of certain determinants in these populations, the models were adapted. In the models for persons of Moroccan and Turkish origin, migration background was excluded and the highest income levels were merged. Being a student was merged with unknown/other socioeconomic position in all models on the population aged ≥60 years.

In all four populations of non-Dutch origin, the odds of not having received an autumn 2022 booster increased as education level decreased (Table 4). This association was strongest among persons of Moroccan and Turkish origin, with aOR=4.41 and aOR=3.65, respectively, for primary education compared to master/doctor level. Living in highly or extremely urban areas compared to a rural area were associated with lower uptake in all populations of non-Dutch origin (aORs between 1.15 and 3.53), while living in somewhat or moderately urban areas were only associated with lower uptake in the populations of Moroccan and Turkish origin (aORs between 1.53 and 2.50). This was in contrast to the effect of urbanisation level with being unvaccinated in the population of Dutch origin (aORs between 0.81 and 0.86). Likewise, compared to high medical risk, no medical risk was associated with lower uptake (aORs between 1.12 and 1.55), but for moderate medical risk this association was only found in the populations of Moroccan and Turkish origin (aOR=1.35 and aOR=1.23, respectively). The odds of not having received an autumn 2022 booster increased as income declined, but this was not as explicit in the population of Turkish origin. Compared to working in employment, being a pensioner (aORs between 0.47 and 0.73) was associated with higher uptake in all populations of non-Dutch origin, while being on benefits (aORs between 0.75 and 0.89) and other/unknown socioeconomic position (aORs between 0.66 and 0.81) were associated with higher uptake among persons of Dutch-Caribbean and Surinamese origin only. Being self-employed was associated with lower uptake in population of Surinamese origin (aOR=1.20). Compared to age ≥80 years, being aged 60-69 years was associated with lower uptake in all populations of non-Dutch origin, while the association with being aged 70-79 varied (Moroccan: aOR=0.79, Turkish: aOR=1.13, Dutch-Caribbean and Surinamese: no significant association). Finally, being female was associated with lower uptake of the autumn 2022 booster, especially among persons of Moroccan origin (aOR=2.00). Associations of age, sex, income, education level, socioeconomic position and medical risk in the population of Dutch origin were largely comparable with those among persons of Dutch-Caribbean and Surinamese origin.

**Table 4.**
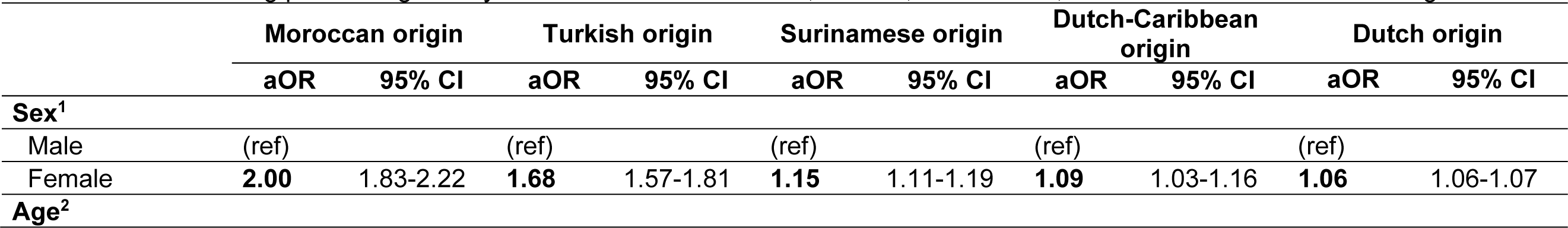
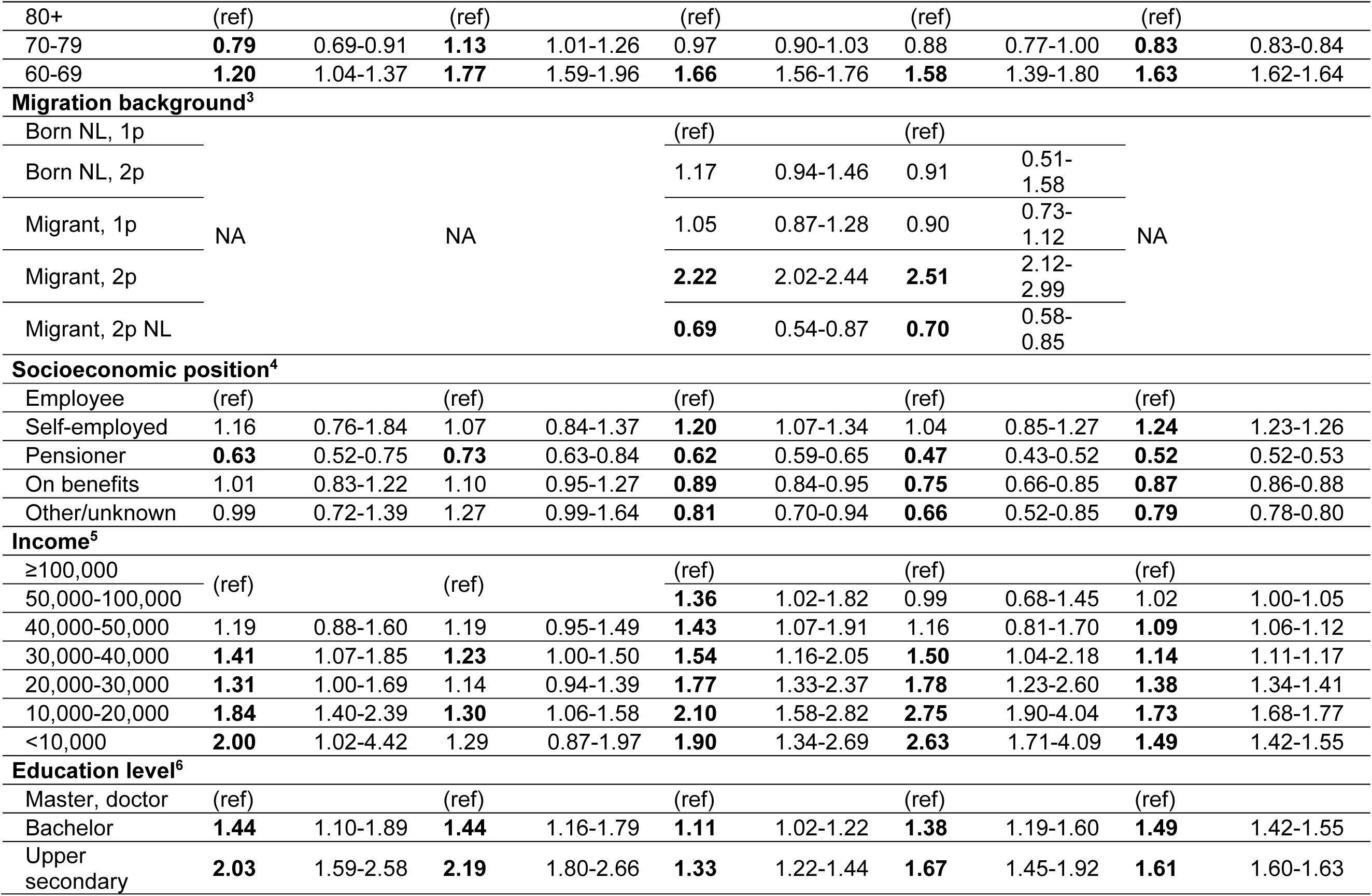
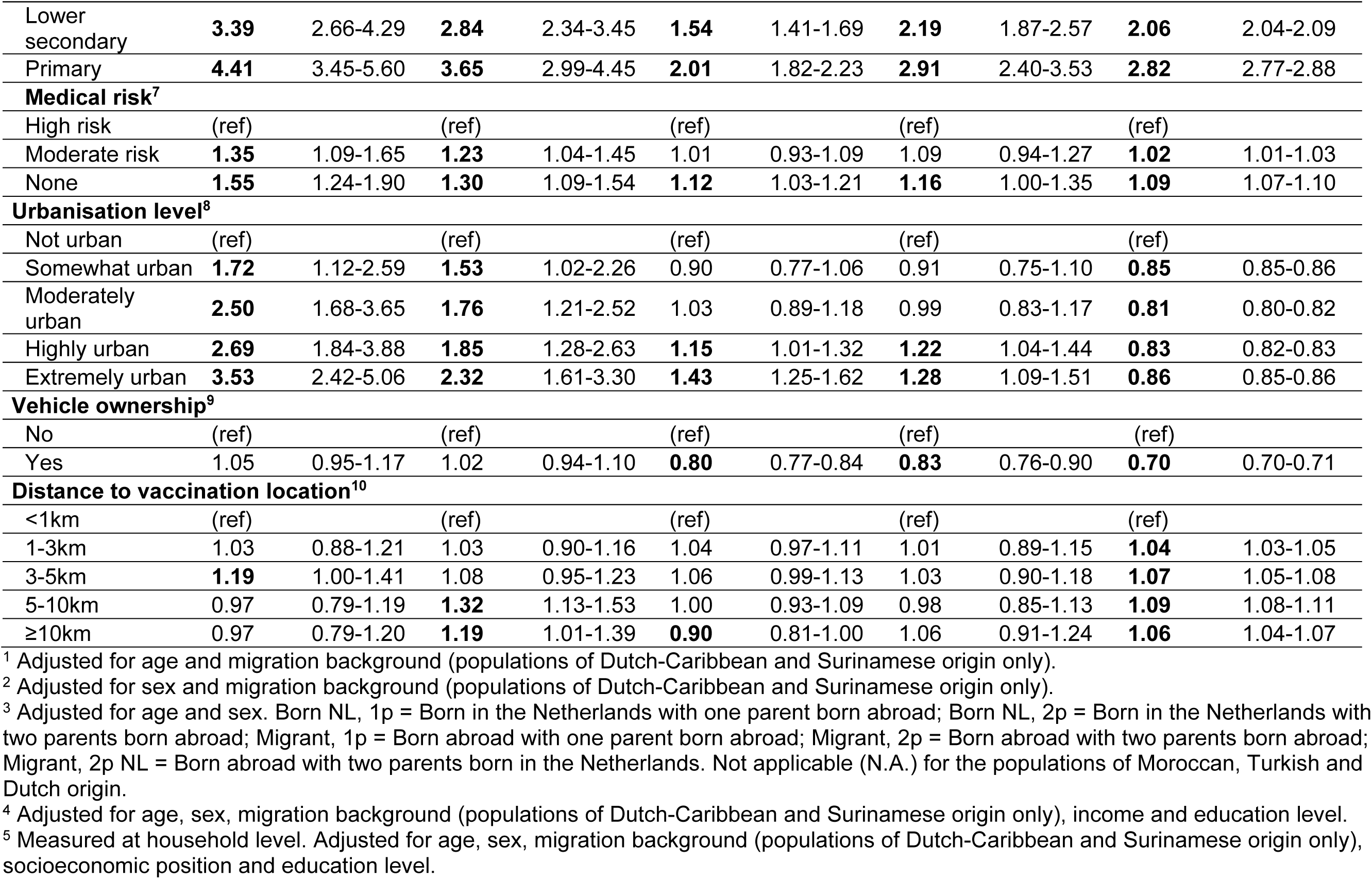
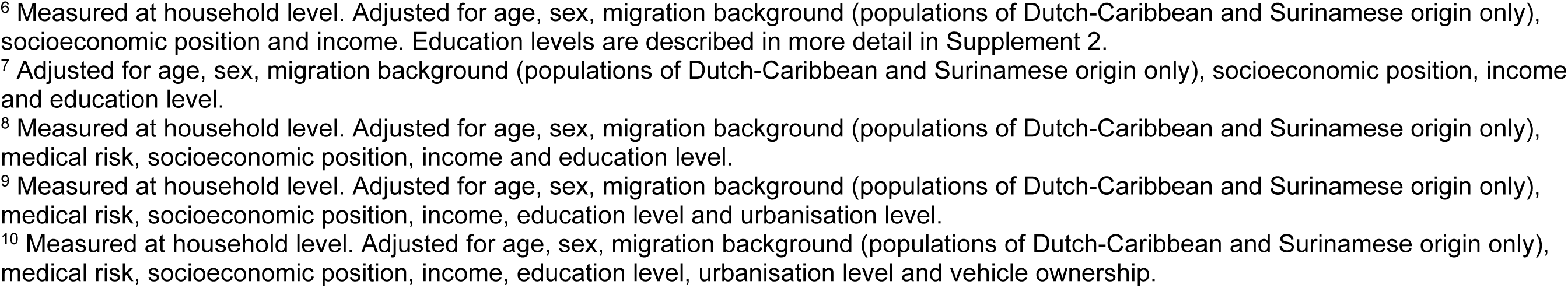
Results of multilevel multivariable models of the association between determinants and being unvaccinated in the autumn 2022 COVID-19 booster round among persons aged 60 years and older of Moroccan, Turkish, Surinamese, Dutch-Caribbean and Dutch origin.

|  | Moroccan origin |  | Turkish origin |  | Surinamese origin |  | Dutch-Caribbean origin |  | Dutch origin |  |
| --- | --- | --- | --- | --- | --- | --- | --- | --- | --- | --- |
|  | aOR | 95% CI | aOR | 95% CI | aOR | 95% CI | aOR | 95% CI | aOR | 95% CI |
| **Sex<sup>1</sup>** |  |  |  |  |  |  |  |  |  |  |
| Male | (ref) |  | (ref) |  | (ref) |  | (ref) |  | (ref) |  |
| Female | **2.00** | 1.83-2.22 | **1.68** | 1.57-1.81 | **1.15** | 1.11-1.19 | **1.09** | 1.03-1.16 | **1.06** | 1.06-1.07 |
| **Age<sup>2</sup>** |  |  |  |  |  |  |  |  |  |  |
| 80+ | (ref) |  | (ref) |  | (ref) |  | (ref) |  | (ref) |  |
| 70-79 | **0.79** | 0.69-0.91 | **1.13** | 1.01-1.26 | 0.97 | 0.90-1.03 | 0.88 | 0.77-1.00 | **0.83** | 0.83-0.84 |
| 60-69 | **1.20** | 1.04-1.37 | **1.77** | 1.59-1.96 | **1.66** | 1.56-1.76 | **1.58** | 1.39-1.80 | **1.63** | 1.62-1.64 |
| **Migration background<sup>3</sup>** |  |  |  |  |  |  |  |  |  |  |
| Born NL, 1p |  |  |  |  | (ref) |  | (ref) |  |  |  |
| Born NL, 2p |  |  |  |  | 1.17 | 0.94-1.46 | 0.91 | 0.51-1.58 |  |  |
| Migrant, 1p | NA |  | NA |  | 1.05 | 0.87-1.28 | 0.90 | 0.73-1.12 | NA |  |
| Migrant, 2p |  |  |  |  | **2.22** | 2.02-2.44 | **2.51** | 2.12-2.99 |  |  |
| Migrant, 2p NL |  |  |  |  | **0.69** | 0.54-0.87 | **0.70** | 0.58-0.85 |  |  |
| **Socioeconomic position<sup>4</sup>** |  |  |  |  |  |  |  |  |  |  |
| Employee | (ref) |  | (ref) |  | (ref) |  | (ref) |  | (ref) |  |
| Self-employed | 1.16 | 0.76-1.84 | 1.07 | 0.84-1.37 | **1.20** | 1.07-1.34 | 1.04 | 0.85-1.27 | **1.24** | 1.23-1.26 |
| Pensioner | **0.63** | 0.52-0.75 | **0.73** | 0.63-0.84 | **0.62** | 0.59-0.65 | **0.47** | 0.43-0.52 | **0.52** | 0.52-0.53 |
| On benefits | 1.01 | 0.83-1.22 | 1.10 | 0.95-1.27 | **0.89** | 0.84-0.95 | **0.75** | 0.66-0.85 | **0.87** | 0.86-0.88 |
| Other/unknown | 0.99 | 0.72-1.39 | 1.27 | 0.99-1.64 | **0.81** | 0.70-0.94 | **0.66** | 0.52-0.85 | **0.79** | 0.78-0.80 |
| **Income<sup>5</sup>** |  |  |  |  |  |  |  |  |  |  |
| ≥100,000 | (ref) |  | (ref) |  | (ref) |  | (ref) |  | (ref) |  |
| 50,000-100,000 |  |  |  |  | **1.36** | 1.02-1.82 | 0.99 | 0.68-1.45 | 1.02 | 1.00-1.05 |
| 40,000-50,000 | 1.19 | 0.88-1.60 | 1.19 | 0.95-1.49 | **1.43** | 1.07-1.91 | 1.16 | 0.81-1.70 | **1.09** | 1.06-1.12 |
| 30,000-40,000 | **1.41** | 1.07-1.85 | **1.23** | 1.00-1.50 | **1.54** | 1.16-2.05 | **1.50** | 1.04-2.18 | **1.14** | 1.11-1.17 |
| 20,000-30,000 | **1.31** | 1.00-1.69 | 1.14 | 0.94-1.39 | **1.77** | 1.33-2.37 | **1.78** | 1.23-2.60 | **1.38** | 1.34-1.41 |
| 10,000-20,000 | **1.84** | 1.40-2.39 | **1.30** | 1.06-1.58 | **2.10** | 1.58-2.82 | **2.75** | 1.90-4.04 | **1.73** | 1.68-1.77 |
| <10,000 | **2.00** | 1.02-4.42 | 1.29 | 0.87-1.97 | **1.90** | 1.34-2.69 | **2.63** | 1.71-4.09 | **1.49** | 1.42-1.55 |
| **Education level<sup>6</sup>** |  |  |  |  |  |  |  |  |  |  |
| Master, doctor | (ref) |  | (ref) |  | (ref) |  | (ref) |  | (ref) |  |
| Bachelor | **1.44** | 1.10-1.89 | **1.44** | 1.16-1.79 | **1.11** | 1.02-1.22 | **1.38** | 1.19-1.60 | **1.49** | 1.42-1.55 |
| Upper secondary | **2.03** | 1.59-2.58 | **2.19** | 1.80-2.66 | **1.33** | 1.22-1.44 | **1.67** | 1.45-1.92 | **1.61** | 1.60-1.63 |
| Lower secondary | **3.39** | 2.66-4.29 | **2.84** | 2.34-3.45 | **1.54** | 1.41-1.69 | **2.19** | 1.87-2.57 | **2.06** | 2.04-2.09 |
| Primary | **4.41** | 3.45-5.60 | **3.65** | 2.99-4.45 | **2.01** | 1.82-2.23 | **2.91** | 2.40-3.53 | **2.82** | 2.77-2.88 |
| **Medical risk<sup>7</sup>** |  |  |  |  |  |  |  |  |  |  |
| High risk | (ref) |  | (ref) |  | (ref) |  | (ref) |  | (ref) |  |
| Moderate risk | **1.35** | 1.09-1.65 | **1.23** | 1.04-1.45 | 1.01 | 0.93-1.09 | 1.09 | 0.94-1.27 | **1.02** | 1.01-1.03 |
| None | **1.55** | 1.24-1.90 | **1.30** | 1.09-1.54 | **1.12** | 1.03-1.21 | **1.16** | 1.00-1.35 | **1.09** | 1.07-1.10 |
| **Urbanisation level<sup>8</sup>** |  |  |  |  |  |  |  |  |  |  |
| Not urban | (ref) |  | (ref) |  | (ref) |  | (ref) |  | (ref) |  |
| Somewhat urban | **1.72** | 1.12-2.59 | **1.53** | 1.02-2.26 | 0.90 | 0.77-1.06 | 0.91 | 0.75-1.10 | **0.85** | 0.85-0.86 |
| Moderately urban | **2.50** | 1.68-3.65 | **1.76** | 1.21-2.52 | 1.03 | 0.89-1.18 | 0.99 | 0.83-1.17 | **0.81** | 0.80-0.82 |
| Highly urban | **2.69** | 1.84-3.88 | **1.85** | 1.28-2.63 | **1.15** | 1.01-1.32 | **1.22** | 1.04-1.44 | **0.83** | 0.82-0.83 |
| Extremely urban | **3.53** | 2.42-5.06 | **2.32** | 1.61-3.30 | **1.43** | 1.25-1.62 | **1.28** | 1.09-1.51 | **0.86** | 0.85-0.86 |
| **Vehicle ownership<sup>9</sup>** |  |  |  |  |  |  |  |  |  |  |
| No | (ref) |  | (ref) |  | (ref) |  | (ref) |  | (ref) |  |
| Yes | 1.05 | 0.95-1.17 | 1.02 | 0.94-1.10 | **0.80** | 0.77-0.84 | **0.83** | 0.76-0.90 | **0.70** | 0.70-0.71 |
| **Distance to vaccination location<sup>10</sup>** |  |  |  |  |  |  |  |  |  |  |
| <1km | (ref) |  | (ref) |  | (ref) |  | (ref) |  | (ref) |  |
| 1-3km | 1.03 | 0.88-1.21 | 1.03 | 0.90-1.16 | 1.04 | 0.97-1.11 | 1.01 | 0.89-1.15 | **1.04** | 1.03-1.05 |
| 3-5km | **1.19** | 1.00-1.41 | 1.08 | 0.95-1.23 | 1.06 | 0.99-1.13 | 1.03 | 0.90-1.18 | **1.07** | 1.05-1.08 |
| 5-10km | 0.97 | 0.79-1.19 | **1.32** | 1.13-1.53 | 1.00 | 0.93-1.09 | 0.98 | 0.85-1.13 | **1.09** | 1.08-1.11 |
| ≥10km | 0.97 | 0.79-1.20 | **1.19** | 1.01-1.39 | **0.90** | 0.81-1.00 | 1.06 | 0.91-1.24 | **1.06** | 1.04-1.07 |

<sup>1</sup> Adjusted for age and migration background (populations of Dutch-Caribbean and Surinamese origin only).
<sup>2</sup> Adjusted for sex and migration background (populations of Dutch-Caribbean and Surinamese origin only).
<sup>3</sup> Adjusted for age and sex. Born NL, 1p = Born in the Netherlands with one parent born abroad; Born NL, 2p = Born in the Netherlands with two parents born abroad; Migrant, 1p = Born abroad with one parent born abroad; Migrant, 2p = Born abroad with two parents born abroad; Migrant, 2p NL = Born abroad with two parents born in the Netherlands. Not applicable (N.A.) for the populations of Moroccan, Turkish and Dutch origin.
<sup>4</sup> Adjusted for age, sex, migration background (populations of Dutch-Caribbean and Surinamese origin only), income and education level.
<sup>5</sup> Measured at household level. Adjusted for age, sex, migration background (populations of Dutch-Caribbean and Surinamese origin only), socioeconomic position and education level.
<sup>6</sup> Measured at household level. Adjusted for age, sex, migration background (populations of Dutch-Caribbean and Surinamese origin only), socioeconomic position and income. Education levels are described in more detail in Supplement 2.
<sup>7</sup> Adjusted for age, sex, migration background (populations of Dutch-Caribbean and Surinamese origin only), socioeconomic position, income and education level.
<sup>8</sup> Measured at household level. Adjusted for age, sex, migration background (populations of Dutch-Caribbean and Surinamese origin only), medical risk, socioeconomic position, income and education level.
<sup>9</sup> Measured at household level. Adjusted for age, sex, migration background (populations of Dutch-Caribbean and Surinamese origin only), medical risk, socioeconomic position, income, education level and urbanisation level.
<sup>10</sup> Measured at household level. Adjusted for age, sex, migration background (populations of Dutch-Caribbean and Surinamese origin only), medical risk, socioeconomic position, income, education level, urbanisation level and vehicle ownership.

Vehicle ownership was only associated with lower odds of not having received an autumn 2022 booster in the populations of Dutch-Caribbean, Surinamese (and Dutch) origin. The associations between not having received an autumn 2022 booster and distance to vaccination location were ambiguous and varied between subgroups.

Being a migrant with two foreign-born parents, compared to being born in the Netherlands with one parent born abroad, was associated with higher odds of not having received an autumn 2022 booster in the populations of Dutch-Caribbean and Surinamese origin (the only populations for which migration background was included) (aOR=2.51 and aOR=2.22, respectively). Being a migrant with two parents born in the Netherlands was associated with higher uptake in these populations (aOR=0.70 and aOR=0.69, respectively). The effect of being a migrant with two foreign-born parents decreased after adding socioeconomic position, income and education level to the model for persons of Dutch-Caribbean origin and, albeit less strongly, for persons of Surinamese origin (Supplement 5 Table S5.2). This indicates that socioeconomic status partially mediated the effect of being a migrant with two foreign-born parents on being unvaccinated in the autumn 2022 booster round. The associations weakened slightly after adding urbanisation level as well, indicating that this determinant might also have a mediating effect.

### 3.4 Population attributable fractions (PAFs) of determinants of being unvaccinated in the primary vaccination round

The PAFs are presented in Figure 1. The determinants with highest PAFs for being unvaccinated against COVID-19 in the primary vaccination round among persons aged ≥18 years of Dutch-Caribbean, Moroccan, Surinamese and Turkish origin were being aged 18-35 years (PAFs between 20.8% and 28.4%), being born in the Netherlands to two foreign-born parents (PAFs between 10.7% and 19.9%), being a migrant with two foreign-born parents (PAFs between 8.5% and 36.1%), not belonging to a medical risk group (PAFs between 16.4% and 22.8%) and having an annual household income below €30,000 (PAFs between 8.3% and 19.5%). The importance of these determinants in terms of PAF varied by country of origin. The impact of being aged 18-35 years and being a migrant with two foreign-born parents were particularly large among persons of Dutch-Caribbean origin (PAF=28.4% and PAF=36.1%, respectively). The PAFs for low income were largest in the populations of Dutch-Caribbean and Surinamese origin.

**Figure 1.**
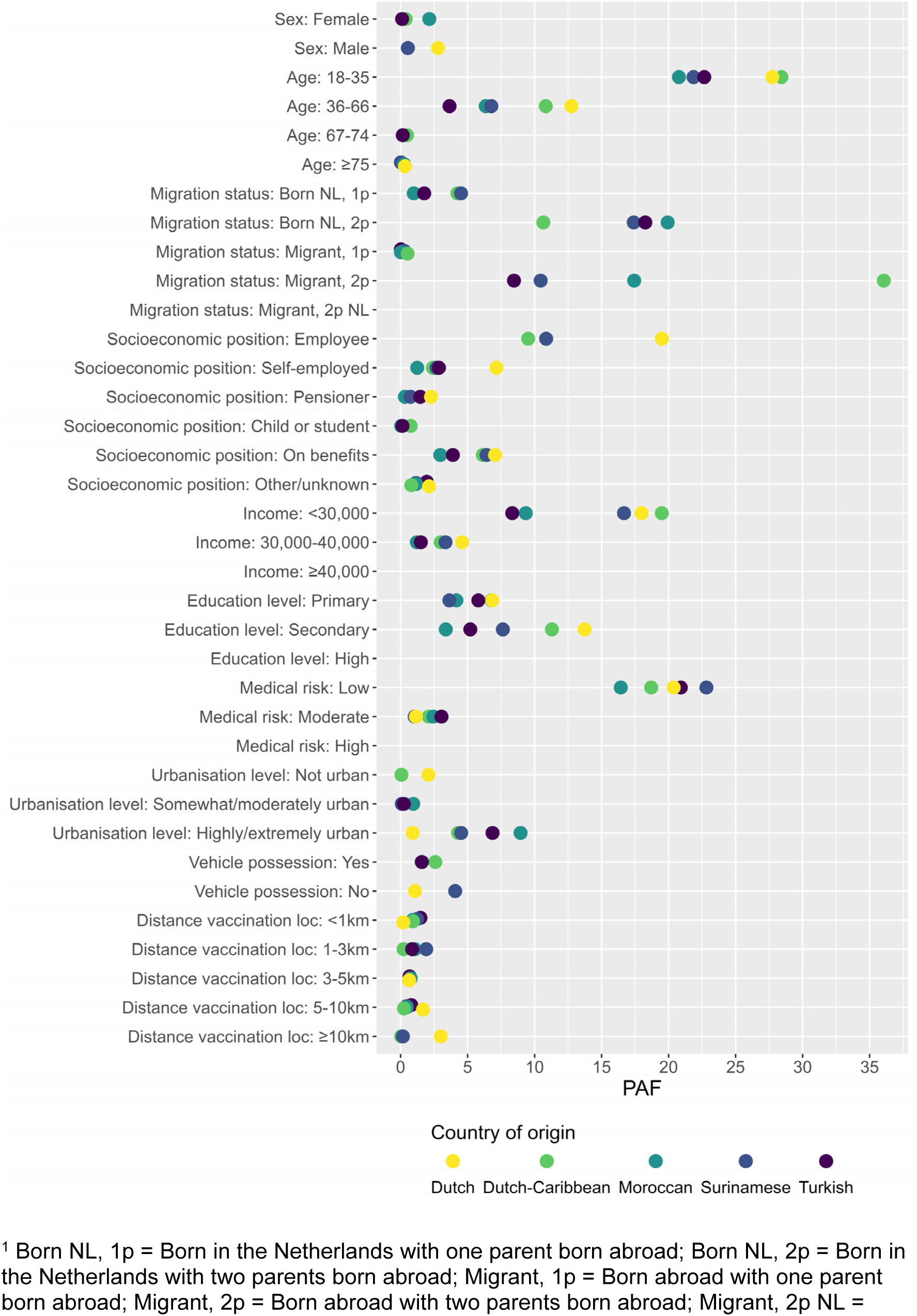
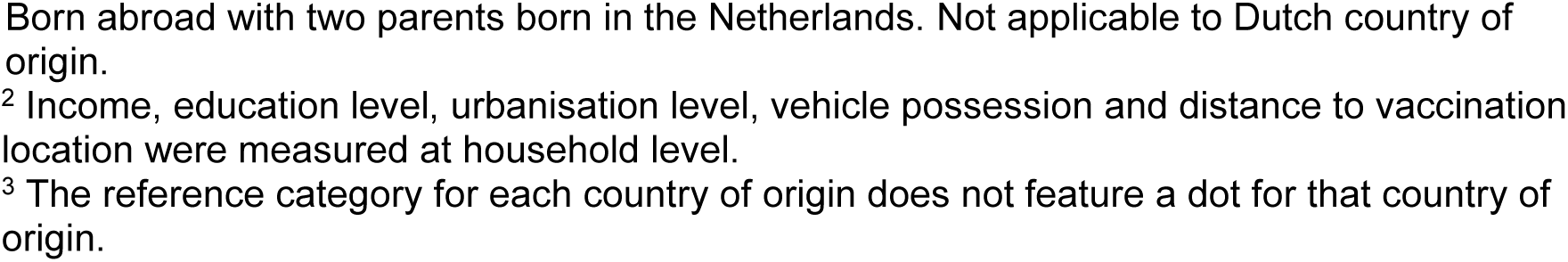
Population attributable fractions (PAFs) of the contribution of each determinant to being unvaccinated in the primary COVID-19 vaccination round by country of origin among persons aged 18 years and older.

Some high PAFs were observed in specific populations. Among persons of Dutch-Caribbean origin, age 36-66 (PAF=10.8%) contributed strongly to being unvaccinated in the primary vaccination round. The PAFs of being an employee, compared to a pensioner (Dutch-Caribbean origin) or student (Surinamese origin), were quite high due to the large number of individuals working in employment (PAF=9.5% and PAF=10.9%, respectively). The impact of education level on being unvaccinated was also considerable, particularly completing secondary education versus higher education among persons with of Dutch-Caribbean origin (PAF=11.3%). For the population of Dutch origin, PAFs per determinant were comparable to those of Dutch-Caribbean origin, except for migration background (by definition). Among persons of Moroccan origin, living in a highly or extremely urbanised area had a considerable impact on being unvaccinated (PAF=9.0%).

### 3.5 PAFs of determinants of being unvaccinated in the autumn 2022 booster round for individuals aged 60 years and older

The PAFs of the studied determinants differed considerably between population countries of origin (Figure 2). Only living in highly or extremely urban areas (PAFs between 8.0% and 12.7%) and to a lesser degree primary education level (PAFs between 6.9% and 11.7%) had an impact in all populations of non-Dutch origin. These determinants yielded the only two substantial PAFs for the populations of Moroccan and Turkish origin. In the populations of Dutch-Caribbean and Surinamese origin, the effect of being a migrant and having two foreign-born parents could be investigated and had an important impact (PAF=30.3% and PAF=35.7% respectively). In the population of Dutch origin, primary and secondary education level, compared to high education level, had PAFs of 8.3% and 9.3%, respectively, while living in highly or extremely urban areas had no impact. Younger age (60-69 years) contributed to being unvaccinated among persons of Dutch-Caribbean, Surinamese and Dutch origin (PAFs between 10.6 and 17.0%). Income below €30,000 only had considerable PAFs for the populations of Dutch-Caribbean and Dutch origin (PAF=13.7 and PAF=8.7%, respectively). Low medical risk and, especially among the populations of Moroccan and Turkish origin, vehicle ownership had no impact on being unvaccinated in the autumn 2022 booster round, while they did in the primary vaccination round.

**Figure 2.**
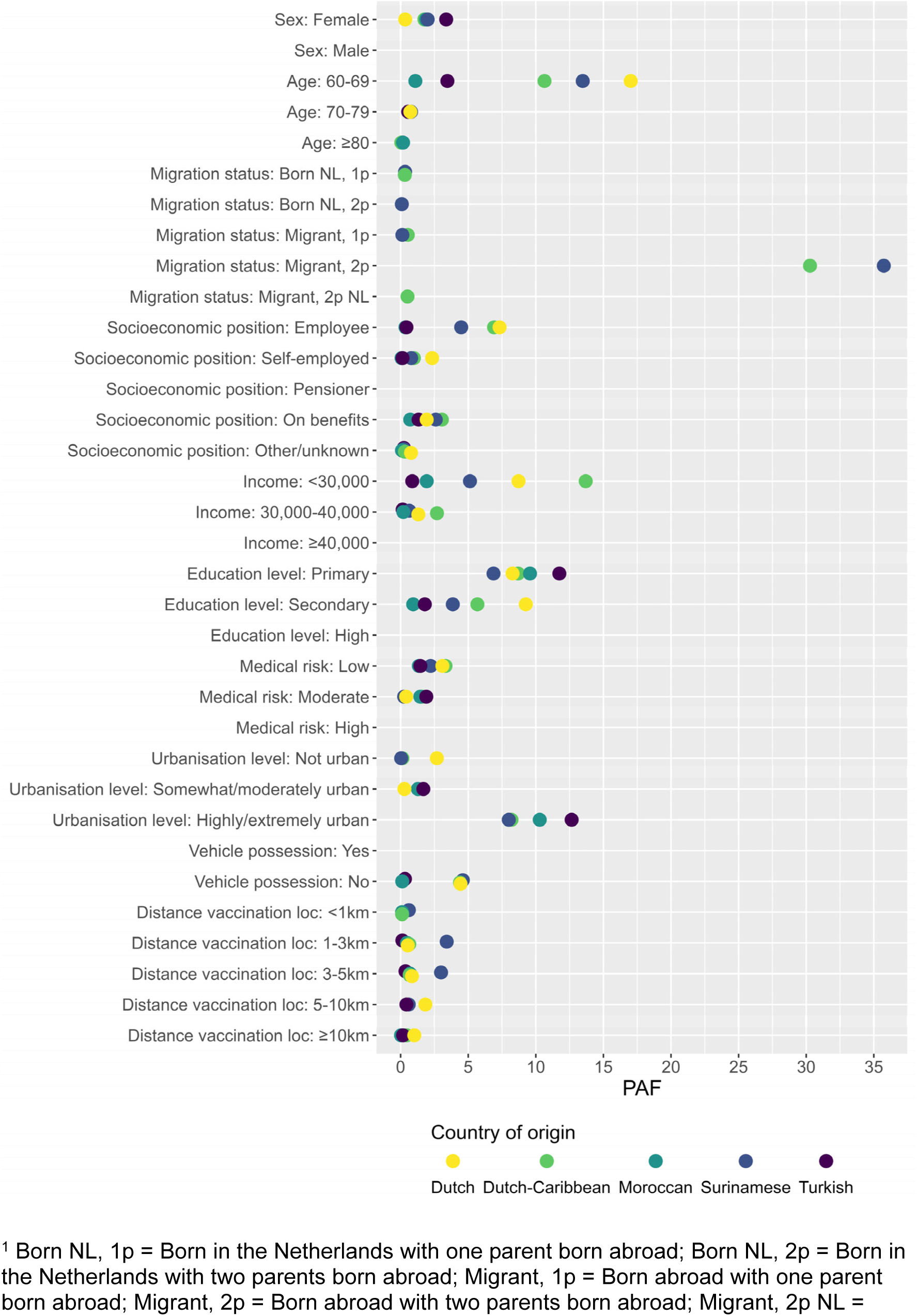
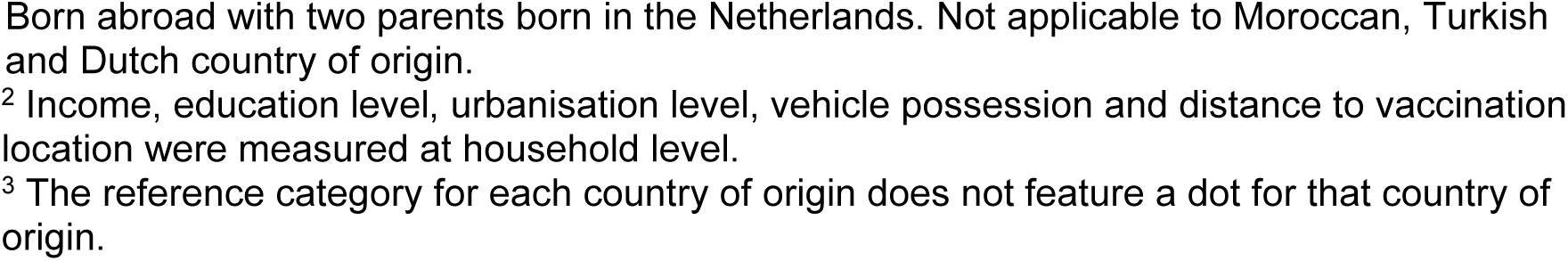
Population attributable fractions (PAFs) of the contribution of each determinant to being unvaccinated in the autumn 2022 COVID-19 booster round by country of origin among persons aged 60 years and older.

## 4. Discussion

We identified determinants associated with being unvaccinated against COVID-19 in the primary and 2022 autumn booster vaccination campaigns among the main populations of non-Dutch origin in the Netherlands: being a migrant with two foreign-born parents, younger age, living in highly or extremely urban areas and having a lower income, lower education level and low medical risk. Being born in the Netherlands with two foreign-born parents, being self-employed, a pensioner, on benefits or having other/unknown socioeconomic position and having a moderate medical risk were additional risk factors for being unvaccinated in the primary vaccination round, while female sex was an additional risk factor in the 2022 autumn booster round in the study population aged ≥60 years. The PAFs largely confirmed the impact of these determinants. We also found differences in the (strength of) determinants of being unvaccinated between persons of Moroccan, Turkish, Surinamese and Dutch-Caribbean origin for both vaccination rounds. This emphasises the importance of tailoring efforts towards increasing vaccine uptake to maximise the impact of vaccination programmes across the whole population of the Netherlands.

The role of age, income and education level in all four populations of non-Dutch origin under study are consistent with previous research, including among migrants and ethnic minorities [2, 11, 20, 23]. In contrast to our findings, belonging to a medical risk group was not an important predictor of being unvaccinated in two previous Dutch studies, which used similar data and risk groups [2, 11]. It is possible that age differences explain the effect of medical risk group and that there was residual confounding by age. Results of studies in other countries are inconsistent [24-26]. The association between being a pensioner and not being vaccinated in the primary vaccination round among the population aged ≥18 years may also be explained by a correlation with age, although similar associations were found in most populations of non-Dutch origin among persons aged ≥60 years.

Being a migrant with two foreign-born parents, compared to being born in the Netherlands with one parent born abroad, was a strong risk factor for being unvaccinated. Previous studies find divergent effects of country of origin and migrant generation on vaccine uptake [27, 28]. We explored to what extent other determinants explained the association between migration background and being unvaccinated. While socioeconomic determinants, and for the autumn booster also urbanisation level, did reduce the odds of being unvaccinated, the association with being a migrant with two foreign-born parents persisted. This suggests that other factors play a role. For instance, social integration, which may be lower for migrants with two foreign-born parents, has been found to be positively associated with higher vaccine acceptance [29, 30].

Determinants that were associated with being unvaccinated in our study were for a considerable part comparable between persons of non-Dutch and Dutch origin. This underlines the need to assess the role of other factors, including religion, trust in authorities, trust in science, social influences and language skills [20, 31, 32], which may also interact with determinants we identified [33]. Survey data and qualitative research might shed more light on these factors. A recent survey-based study by the CBS identified religion, trust in the political and healthcare institutions, social participation and self-reported health as COVID-19 vaccination determinants [34]. It would be relevant to stratify such studies by migration background.

Additionally, studying the ‘transmission’ of vaccine uptake within social networks, as well as how vaccine uptake in these networks influences the decision to get an additional booster each round, seems a promising avenue for further research [35]. Particularly within cities, communities of people with a migration background are often concentrated in certain neighbourhoods, and vaccine hesitancy may easily spread between community members, resulting in lower vaccine uptake in these communities [31, 36-38]. This potentially explains the observed association between living in a highly/extremely urbanised area and lower uptake, which was particularly strong among persons of Moroccan origin. A better understanding of these dynamics may provide the basis for community-based interventions to address vaccine hesitancy. Studies could explore the role of trusted community-based organisations and key figures in the community [33, 39, 40]. Considering the important role of friends and family in reinforcing or alleviating vaccine hesitancy and ultimately in vaccine uptake, studies focused on how vaccine recipients can be ambassadors to recruit others to get vaccinated are needed as well [31, 36, 37].

The observed differences in the influence of some determinants between populations from different countries of origin further highlights the need for community-based interventions targeted at specific ethnic minority and migrant populations. Besides urbanisation level, we observed differences in the influence of education level and income in both vaccination rounds. When targeting interventions, it is relevant to consider both the strength of the associations and the size of the population that they apply to. PAFs can be interpreted as the percentage of not receiving vaccination against COVID-19 attributable to each determinant. The PAFs highlight that groups of young age, migrants with two foreign-born parents, low income, low education level or low medical risk and, in the autumn 2022 booster round, groups living in a highly or extremely urbanised area at population level contributed considerably to low vaccine uptake.

A key strength of this study is the use of population-wide individual-level and household-level data, which avoids ecological fallacies that may result from using aggregated data. By studying the primary vaccination round and the autumn 2022 booster round and four populations of non-Dutch origin as well as people of Dutch origin, we made a comprehensive assessment of determinants of being unvaccinated both initially and for repeat vaccinations.

There are some limitations that need to be considered. Firstly, vaccine uptake was underestimated in the primary vaccination round, as people who did not consent to nationally registering their vaccination were included as unvaccinated. This may have biased the estimates in different ways, depending on the characteristics of persons with higher and lower consent percentages. This is less relevant for the booster vaccination, as this consent rate was very high (99.2%). Secondly, as we used population-level data, we could not include some potential determinants of vaccine uptake, like religion, trust in authorities and science and language skills, which may be especially relevant in migrant and ethnic minority populations [20, 31, 32]. Thirdly, some underlying medical conditions and distance to vaccination locations were hard to identify based on available data, so some misclassification of these determinants may be present.

## 5. Conclusion

This study identified several common risk factors for being unvaccinated against COVID-19 observed in four large populations of non-Dutch origin and found substantial differences in the (strength of the) determinants and corresponding PAFs between the populations of Moroccan, Turkish, Surinamese and Dutch-Caribbean origin. Importantly, the higher odds of being unvaccinated for migrants with two foreign-born parents compared to persons born in the Netherlands with one foreign-born parent, could only partially be explained by socioeconomic variables. This indicates that socioeconomic status only partially mediated this effect and illustrates that interventions specifically targeted at these ethnic minority and migrant populations are needed to optimise the impact of vaccination programmes across the whole population of the Netherlands. Although these findings provide some guidance to target vaccination programmes, qualitative and survey-based research is needed to further understand reasons behind not receiving vaccination in these populations and design (community-based) interventions.

## Data availability

Results are based on non-public microdata from Statistics Netherlands. Under certain conditions, these microdata are accessible for statistical and scientific research. For further information.

## Acknowledgements

We would like to thank Ben Bom for compiling the vaccination location data and William Schuch for helping to prepare these data for analysis. We would like to thank Hendriek Boshuizen and Jan van de Kassteele for their assistance with the statistical analyses.

## Financial support

This research received no specific grant from any funding agency, commercial or not-for-profit sectors.

## Competing interests

The authors declare none.

## Notes

### Competing Interest Statement

The authors have declared no competing interest.

### Author Declarations

The Centre for Clinical Expertise at the National Institute for Public Health and the Environment (the Netherlands) waived ethical approval for this work.

