## Supplement for "Factors associated with lower COVID-19 vaccine uptake among populations with a migration background in the Netherlands"

**Supplementary Material**

Supplement 1: Number of unvaccinated persons and vaccine uptake by country of origin

Table S1 Number of unvaccinated persons and vaccine uptake of at least one COVID-19 vaccination in the primary vaccination round and of the booster in the autumn 2022 round by country of origin.

|  | **≥1 COVID-19 vaccination in the primary vaccination round** | | | **Booster in the autumn 2022 round** | |
| --- | --- | --- | --- | --- | --- |
| **Country or region of origin^1^** | **N unvaccinated^2^** | **Vaccine uptake (%)** | **N unvaccinated^3^** | | **Vaccine uptake (%)^3^** |
| The Netherlands | 1,614,910 | 84.9 | 1,049,030 | | 69.5 |
| Morocco | 175,280 | 40.5 | 24,360 | | 8.3 |
| Middle and Eastern European countries within the EU**^2^** | 167,080 | 45.1 | 9,060 | | 51.4 |
| Turkey | 155,210 | 53.4 | 23,180 | | 14.7 |
| Other Asia**^2^** | 139,320 | 72.2 | 28,240 | | 38.7 |
| Suriname | 112,970 | 62.9 | 29,340 | | 46.4 |
| Other countries of the EU**^2^** | 108,360 | 79.0 | 56,340 | | 69.1 |
| Other Africa**^2^** | 93,030 | 59.2 | 12,500 | | 35.7 |
| Caribbean part of the Kingdom of the Netherlands | 65,720 | 51.8 | 8,350 | | 50.6 |
| Indonesia | 59,210 | 83.2 | 41,420 | | 68.7 |
| Other America/Oceania**^2^** | 52,830 | 72.9 | 8,370 | | 59.2 |
| Other European countries**^2^** | 46,320 | 71.0 | 11,060 | | 56.9 |
| GIPS-countries**^2,4^** | 45,590 | 68.0 | 6,430 | | 53.1 |
| Former or associated member states of the Commonwealth of Independent States**^2^** | 17,020 | 63.1 | 2,460 | | 49.4 |

*Note.* Following CBS publication guidelines, all tables exclude frequencies below ten and all numbers and percentages are rounded to the nearest ten to avoid personally identifiable information.

^1^ Countries or regions of origin were ranked in descending order in this column, i.e. in descending order of the number of persons who have not received any COVID-19 vaccination. The total study population consisted of the population of the Netherlands aged 18 years and older.

^2^ Considering the heterogeneity in the population in different countries in this region, and hereby limited implications for policy and practice, large, broadly defined regions of origin were excluded.

^3^ The total study population consisted of people aged 60 years and older with at least one previous COVID-19 vaccination.

^4^ Greece, Italy, Portugal, Spain.

Supplement 2: Overview of the determinants

Table S2 Overview of the determinants and data sources.

| **Determinant** | **Level** | **Reference date** | | **Data-base** |
| --- | --- | --- | --- | --- |
|  |  | **Primary vaccination round** | **Autumn 2022 booster round** |  |
| Vaccine uptake | Yes; No (or no informed consent) | 05-01-2021 – 18-11-2021 | 19-09-2022 – 15-09-2023 | CIMS |
| **Individual level** | | | | |
| Sex | Male; female | 05-01-2021 | 19-09-2022 | CBS |
| Age | 18-22; 23-27; 28-32; 33-37; 38-42; 43-47; 48-52; 53-57; 58-62; 63-67; 68-74; ≥75 | 05-01-2021 | 19-09-2022 | CBS |
| Migration background^1^ | The Netherlands; Born in the Netherlands with one parent born abroad; Born in the Netherlands with two parents born abroad; Born abroad with one parent born abroad; Born abroad with two parents born abroad; Born abroad with two parents born in the Netherlands | 2021 | 2023 | CBS |
| Socioeconomic position^2^ | In employment; Self-employed; (School)child or Student, Pensioner; On benefits; Other/Unknown | 2021 | 2022 | CBS |
| Medical risk^3^ | Low medical risk; Moderate medical risk; High medical risk | 2020^4^ | 2021^4^ | Vektis |
| **Household level** | | | | |
| Income^5^ | <10,000; 10,000-20,000; 20,000-30,000; 30,000-40,000; 40,000-50,000; 50,000-100,000; ≥100,000 | 2021 | 2022 | CBS |
| Education level | Primary (basisschool); Lower secondary = prevocational secondary education (vmbo), lower secondary vocational training and assistant’s training (mbo1), the first 3 years of senior general secondary education (havo) and pre-university secondary education (vwo); Upper secondary = basic vocational training (mbo2), vocational training (mbo3), middle management and specialist education (mbo4), complete secondary education (havo/vwo); Bachelor (hbo-/wo-bachelor); Master, doctor (hbo-/wo-master, doctor) | 2021 | 2021 | CBS |
| Urbanisation level | Not urban; Somewhat urban; Moderately urban; Highly urban; Extremely urban; Unknown | 2021 | 2022 | CBS |
| Vehicle ownership | No; Yes | 2021 | 2023 | CBS |
| Distance to closest vaccination location | <1km, 1-3km, 3-5km, 5-10km, ≥10km | 01-07-2021 –  18-11-2021 | 19-09-2022 –  13-11-2022 | CBS; RIVM^6^ |

^1^ If a person was born in the Netherlands but their mother was born abroad, country of origin was defined as the mother's country of birth. If only the father was born abroad, their father's country of birth was the person's country of origin.

^2^ Socioeconomic position refers to an individual’s largest source of income, complemented with the status (School)child or Student. On benefits includes unemployment benefits, social assistance benefits, disability benefits and other benefits.

^3^ Approximated based on healthcare utilisation and medication prescription data, as described by Pijpers et al. [1].

^4^ In case of rare medical conditions, data from 2016-2020 were included.

^5^ Annual disposable household income in euros, standardised by household type/size.

^6^ Distance to the closest long-term vaccination location was estimated as the crow flies, based on XY coordinates of the six-digit postal code where each person’s home address was registered and a log of all vaccination locations according to official appointment booking system (XY coordinates).

Supplement 3: Vaccine uptake of at least one COVID-19 vaccination in the primary vaccination round by country of origin and determinant

Table S3.1 Vaccine uptake of at least one COVID-19 vaccination in the primary vaccination round among persons aged 18 years and older per country of origin by age.

|  | **Moroccan origin** | | **Turkish origin** | | **Surinamese origin** | | **Dutch-Caribbean origin** | | **Dutch origin** | |
| --- | --- | --- | --- | --- | --- | --- | --- | --- | --- | --- |
| **Age** | **N** | **Uptake N (%)** | **N** | **Uptake N (%)** | **N** | **Uptake N (%)** | **N** | **Uptake N (%)** | **N** | **Uptake N (%)** |
| 18-22 | 36.610 | 6.760 (18) | 35.130 | 10.840 (31) | 25.930 | 12.450 (48) | 16.070 | 6.430 (40) | 774,790 | 597.240 (77) |
| 23-27 | 32.090 | 6.100 (19) | 37.540 | 11.940 (32) | 26.350 | 11.190 (42) | 18.260 | 7.000 (38) | 745,180 | 553.330 (74) |
| 28-32 | 32.620 | 7.140 (22) | 39.430 | 15.280 (39) | 29.230 | 12.660 (43) | 17.070 | 6.530 (38) | 750,360 | 550.920 (73) |
| 33-37 | 31.760 | 9.920 (31) | 36.000 | 17.810 (49) | 30.470 | 15.020 (49) | 14.610 | 6.240 (43) | 704,150 | 540.050 (77) |
| 38-42 | 33.070 | 14.010 (42) | 38.130 | 22.660 (59) | 27.100 | 15.570 (57) | 12.640 | 6.320 (50) | 697,920 | 563.620 (81) |
| 43-47 | 31.240 | 15.860 (51) | 35.340 | 22.710 (64) | 25.710 | 17.000 (66) | 10.450 | 5.850 (56) | 740,040 | 621.240 (84) |
| 48-52 | 28.080 | 15.750 (56) | 35.340 | 24.010 (68) | 30.840 | 21.870 (71) | 10.820 | 6.610 (61) | 961,020 | 824.950 (86) |
| 53-57 | 21.630 | 13.140 (61) | 27.990 | 19.560 (70) | 29.480 | 21.970 (75) | 9.380 | 5.990 (64) | 998,150 | 875.750 (88) |
| 58-62 | 15.510 | 9.880 (64) | 18.440 | 13.060 (71) | 26.610 | 21.030 (79) | 9.030 | 6.200 (69) | 968,580 | 873.460 (90) |
| 63-67 | 9.750 | 6.450 (66) | 9.510 | 6.480 (68) | 21.340 | 17.290 (81) | 7.210 | 5.220 (72) | 873,750 | 801.380 (92) |
| 68-74 | 10.250 | 6.750 (66) | 9.860 | 6.490 (66) | 18.030 | 14.700 (82) | 6.570 | 4.940 (75) | 1,159,940 | 1.075.970 (93) |
| 75+ | 11.870 | 7.450 (63) | 10.390 | 7.060 (68) | 13.290 | 10.650 (80) | 4.110 | 3.180 (77) | 1,286,550 | 1.167.620 (91) |

*Note.* Following CBS publication guidelines, all tables exclude frequencies below ten and all numbers and percentages are rounded to the nearest ten to avoid personally identifiable information.

Table S3.2 Vaccine uptake of at least one COVID-19 vaccination in the primary vaccination round among persons aged 18 years and older per country of origin by determinant.

|  | **Moroccan origin** | | **Turkish origin** | | **Surinamese origin** | | **Dutch-Caribbean origin** | | **Dutch origin** | |
| --- | --- | --- | --- | --- | --- | --- | --- | --- | --- | --- |
| **Determinant** | **N** | **Uptake N (%)** | **N** | **Uptake N (%)** | **N** | **Uptake N (%)** | **N** | **Uptake N (%)** | **N** | **Uptake N**  **(%)** |
| **Sex** | | | | | | | | | | |
| Male | 149,160 | 62,630 (42) | 171,810 | 91,590 (53) | 142,550 | 87,970 (62) | 67,850 | 34,880 (51) | 5,278,860 | 4,454,590 (84) |
| Female | 145,320 | 56,580 (39) | 161,290 | 86,290 (54) | 161,820 | 103,420 (64) | 68,380 | 35,630 (52) | 5,381,570 | 4,590,920 (85) |
| **Migration background^1^** | | | | | | | | | | |
| Born NL, 1p | 12,770 | 5,480 (43) | 17,390 | 7,690 (44) | 42,430 | 25,020 (59) | 24,720 | 16,440 (67) | NA | NA |
| Born NL, 2p | 112,310 | 25,300 (23) | 122,620 | 47,110 (38) | 86,370 | 40,120 (46) | 18,430 | 5,890 (32) |  |  |
| Migrant, 1p | 250 | 120 (48) | 480 | 260 (54) | 3,000 | 2,010 (67) | 4,320 | 3,180 (74) |  |  |
| Migrant, 2p | 169,020 | 88,220 (52) | 192,380 | 122,680 (64) | 170,950 | 122,950 (72) | 81,880 | 39,090 (48) |  |  |
| Migrant, 2p NL | 120 | 90 (73) | 220 | 150 (68) | 1,620 | 1,300 (80) | 6,880 | 5,910 (86) |  |  |
| **Socio-economic position** | | | | | | | | | | |
| Employee | 118,740 | 49,730 (42) | 145,180 | 81,260 (56) | 158,730 | 100,920 (64) | 69,540 | 37,880 (54) | 5,322,300 | 4,500,580 (85) |
| Self-employed | 21,950 | 6,830 (31) | 35,630 | 16,520 (46) | 18,210 | 10,420 (57) | 7,960 | 3,810 (48) | 798,650 | 617,180 (77) |
| Pensioner | 28,500 | 17,890 (63) | 26,050 | 17,180 (66) | 41,830 | 33,670 (80) | 13,940 | 10,530 (76) | 2,928,190 | 2,685,300 (92) |
| Child or student | 24,430 | 5,250 (21) | 25,020 | 8,920 (36) | 22,050 | 11,410 (52) | 16,980 | 7,170 (42) | 566,690 | 468,560 (83) |
| On benefits | 76,470 | 30,780 (40) | 75,490 | 42,240 (56) | 53,970 | 30,160 (56) | 23,750 | 9,460 (40) | 272,390 | 207,500 (76) |
| Other/  unknown | 24,410 | 8,740 (36) | 25,730 | 11,750 (46) | 9,580 | 4,820 (50) | 4,060 | 1,660 (41) | 772,210 | 566,400 (73) |
| **Income^2^** | | | | | | | | | | |
| <10,000 | 9,530 | 2,530 (27) | 10,790 | 4,310 (40) | 10,310 | 5,010 (49) | 10,390 | 4,440 (43) | 223,900 | 163,190 (73) |
| 10,000-20,000 | 109,810 | 45,870 (42) | 91,290 | 48,640 (53) | 69,750 | 39,490 (57) | 36,380 | 14,540 (40) | 1,229,840 | 938,890 (76) |
| 20,000-30,000 | 85,020 | 33,090 (39) | 94,230 | 49,240 (52) | 81,600 | 49,060 (60) | 36,770 | 17,360 (47) | 2,815,900 | 2,367,550 (84) |
| 30,000-40,000 | 52,920 | 21,750 (41) | 75,540 | 41,120 (54) | 71,690 | 47,290 (66) | 26,770 | 15,810 (59) | 2,892,640 | 2,489,020 (86) |
| 40,000-50,000 | 22,320 | 9,530 (43) | 35,610 | 20,200 (57) | 40,960 | 28,710 (70) | 13,810 | 9,340 (68) | 1,851,480 | 1,623,860 (88) |
| 50,000-100,000 | 11,730 | 5,560 (47) | 20,510 | 12,350 (60) | 26,250 | 19,580 (75) | 9,990 | 7,810 (78) | 1,489,450 | 1,326,870 (89) |
| ≥100,000 | 740 | 380 (51) | 1,420 | 920 (65) | 1,720 | 1,410 (82) | 910 | 790 (86) | 141,350 | 127,120 (90) |
| Unknown | 2,410 | 490 (20) | 3,700 | 1,090 (29) | 2,100 | 850 (40) | 1,210 | 410 (34) | 15,870 | 9,020 (57) |
| **Education level^2,3^** | | | | | | | | | | |
| Primary | 53,930 | 23,630 (44) | 51,970 | 27,880 (54) | 16,160 | 10,240 (63) | 7,490 | 2,910 (39) | 177,020 | 131,270 (74) |
| Lower secondary | 69,150 | 29,090 (42) | 71,360 | 38,920 (55) | 49,560 | 30,500 (62) | 19,630 | 8,360 (43) | 1,261,790 | 1,042,610 (83) |
| Upper secondary | 108,350 | 39,780 (37) | 135,280 | 69,010 (51) | 143,260 | 86,590 (60) | 64,740 | 30,330 (47) | 5,045,150 | 4,213,340 (84) |
| Bachelor | 38,400 | 15,470 (40) | 46,830 | 25,620 (55) | 59,090 | 38,740 (66) | 25,680 | 16,020 (62) | 2,595,510 | 2,251,700 (87) |
| Master, doctor | 18,330 | 9,350 (51) | 21,160 | 13,660 (65) | 28,320 | 21,170 (75) | 14,060 | 10,860 (77) | 1,382,250 | 1,248,450 (90) |
| Unknown | 6,330 | 1,900 (30) | 6,510 | 2,800 (43) | 7,990 | 4,150 (52) | 4,630 | 2,040 (44) | 198,720 | 158,150 (80) |
| **Medical risk** |  |  |  |  |  |  |  |  |  |  |
| Low | 218,960 | 79,850 (36) | 251,770 | 126,260 (50) | 216,730 | 126,320 (58) | 106,020 | 52,210 (49) | 7,850,350 | 6,554,940 (83) |
| Moderate | 69,930 | 36,220 (52) | 74,000 | 46,680 (63) | 78,530 | 58,270 (74) | 26,560 | 16,000 (60) | 520,770 | 2,232,170 (89) |
| High | 5,600 | 3,140 (56) | 7,320 | 4,940 (67) | 9,110 | 6,810 (75) | 3,650 | 2,310 (63) | 289,310 | 258,410 (89) |
| **Urbanisation level^2^** | | | | | | | | | | |
| Not urban | 3,760 | 1,820 (48) | 4,410 | 2,570 (58) | 7,130 | 4,860 (68) | 5,560 | 3,580 (64) | 360 | 240 (66) |
| Somewhat urban | 12,000 | 5,200 (43) | 15,130 | 8,720 (58) | 13,700 | 9,310 (68) | 8,550 | 5,440 (64) | 2,035,740 | 1,706,750 (84) |
| Moderately urban | 30,800 | 13,460 (44) | 42,690 | 24,320 (57) | 34,000 | 22,650 (67) | 16,030 | 9,500 (59) | 1,937,680 | 1,662,960 (86) |
| Highly urban | 83,460 | 34,800 (42) | 102,890 | 56,080 (55) | 90,980 | 56,810 (62) | 40,650 | 20,950 (52) | 2,061,160 | 1,766,740 (86) |
| Extremely urban | 164,420 | 63,920 (39) | 167,940 | 86,190 (51) | 158,510 | 97,750 (62) | 65,410 | 31,040 (47) | 2,614,900 | 2,210,470 (85) |
| **Vehicle ownership^2^** | | | | | | | | | | |
| Yes | 212,940 | 84,340 (40) | 252,290 | 134,360 (53) | 201,340 | 132,080 (66) | 79,540 | 44,390 (56) | 8,825,180 | 7,555,260 (86) |
| No | 81,540 | 34,860 (43) | 80,800 | 43,520 (54) | 103,030 | 59,310 (58) | 56,690 | 26,120 (46) | 1,835,250 | 1,490,260 (81) |
| **Distance to vaccination location^2^** | | | | | | | | | | |
| <1km | 35,520 | 13,240 (37) | 38,080 | 18,440 (48) | 37,520 | 22,300 (59) | 15,340 | 6,430 (42) | 351,010 | 294,310 (84) |
| 1-3km | 86,440 | 34,490 (40) | 86,080 | 45,890 (53) | 96,840 | 59,200 (61) | 37,410 | 18,890 (50) | 1,756,250 | 1,495,810 (85) |
| 3-5km | 70,010 | 28,260 (40) | 77,560 | 41,460 (53) | 73,550 | 47,170 (64) | 30,310 | 15,410 (51) | 1,571,880 | 1,335,420 (85) |
| 5-10km | 53,800 | 22,110 (41) | 70,600 | 38,090 (54) | 58,690 | 38,090 (65) | 28,120 | 15,800 (56) | 3,204,990 | 2,734,020 (85) |
| ≥10km | 48,720 | 21,110 (43) | 60,780 | 34,000 (56) | 37,780 | 24,640 (65) | 25,050 | 13,990 (56) | 3,776,310 | 3,185,960 (84) |

*Note.* Following CBS publication guidelines, all tables exclude frequencies below ten and all numbers and percentages are rounded to the nearest ten to avoid personally identifiable information.

^1^ Born NL, 1p = Born in the Netherlands with one parent born abroad; Born NL, 2p = Born in the Netherlands with two parents born abroad; Migrant, 1p = Born abroad with one parent born abroad; Migrant, 2p = Born abroad with two parents born abroad; Migrant, 2p NL = Born abroad with two parents born in the Netherlands.

^2^ Measured at household level.

^3^ Education levels are described in more detail in Supplement 2.

Supplement 4: Vaccine uptake of the booster in the autumn round by country of origin and determinant

Table S4 Vaccine uptake of the booster in the autumn 2022 round among persons aged 60 years and older per country of origin by determinant.

|  | **Moroccan origin** | | **Turkish origin** | | **Surinamese origin** | | **Dutch-Caribbean origin** | | **Dutch origin** | |
| --- | --- | --- | --- | --- | --- | --- | --- | --- | --- | --- |
| **Determinant** | **N** | **Uptake N (%)** | **N** | **Uptake N (%)** | **N** | **Uptake N (%)** | **N** | **Uptake N (%)** | **N** | **Uptake N (%)** |
| **Sex** | | | | | | | | | | |
| Male | 14,440 | 1,530 (11) | 13,310 | 2,360 (18) | 24,030 | 11,600 (48) | 7,760 | 4,030 (52) | 1,632,320 | 1,140,780 (70) |
| Female | 12,110 | 660 (5) | 13,870 | 1,640 (12) | 30,720 | 13,800 (45) | 9,140 | 4,530 (50) | 1,802,570 | 1,245,070 (69) |
| **Age** | | | | | | | | | | |
| 60-69 | 14,530 | 1,010 (7) | 15,740 | 1,950 (12) | 35,370 | 14,780 (42) | 10,620 | 4,880 (46) | 1,615,430 | 1,012,160 (63) |
| 70-79 | 8,620 | 900 (10) | 8,510 | 1,470 (17) | 14,840 | 8,160 (55) | 5,160 | 3,040 (59) | 1,253,180 | 959,990 (77) |
| 80+ | 3,400 | 290 (8) | 2,930 | 570 (20) | 4,540 | 2,460 (54) | 1,130 | 640 (57) | 566,280 | 413,700 (73) |
| **Migratie-status^1^** | | | | | | | | | | |
| Born NL, 1p | . | . | 60 | 40 (75) | 2,000 | 1,300 (65) | 590 | 380 (64) | NA | NA |
| Born NL, 2p | . | . | . | . | 390 | 230 (59) | 60 | 40 (67) |  |  |
| Migrant, 1p | 20 | . | 20 | . | 580 | 370 (64) | 970 | 660 (68) |  |  |
| Migrant, 2p | 26,490 | 2,150 (8) | 27,060 | 3,910 (14) | 51,370 | 23,210 (45) | 12,670 | 5,570 (44) |  |  |
| Migrant, 2p NL | 40 | 20 (54) | 40 | 30 (73) | 410 | 290 (71) | 2,610 | 1,910 (73) |  |  |
| **Socio-economic position** | | | | | | | | | | |
| Employee | 2,440 | 230 (9) | 2,630 | 420 (16) | 11,830 | 4,770 (40) | 3,600 | 1,550 (43) | 538,530 | 313,600 (58) |
| Self-employed | 300 | 30 (10) | 680 | 100 (15) | 1,450 | 570 (39) | 480 | 240 (49) | 128,820 | 69,400 (54) |
| Pensioner | 16,980 | 1,580 (9) | 15,830 | 2,660 (17) | 32,020 | 16,650 (52) | 10,030 | 5,760 (57) | 2,517,620 | 1,862,560 (74) |
| On benefits | 5,540 | 290 (5) | 6,830 | 700 (10) | 8,340 | 2,980 (36) | 2,280 | 770 (34) | 166,290 | 91,010 (55) |
| Other/unknown | 1,280 | 60 (5) | 1,210 | 120 (10) | 1,120 | 450 (40) | 510 | 240 (47) | 83,630 | 49,280 (59) |
| **Income^2^** | | | | | | | | | | |
| <10,000 | 190 | 20 (12) | 310 | 60 (20) | 700 | 320 (45) | 480 | 210 (43) | 14,990 | 8,700 (58) |
| 10,000-20,000 | 14,650 | 940 (6) | 13,600 | 1,800 (13) | 13,260 | 5,390 (41) | 4,190 | 1,470 (35) | 311,080 | 188,680 (61) |
| 20,000-30,000 | 7,500 | 720 (10) | 7,720 | 1,160 (15) | 19,070 | 8,850 (46) | 4,890 | 2,400 (49) | 1,368,420 | 943,220 (69) |
| 30,000-40,000 | 2,570 | 270 (10) | 3,190 | 500 (16) | 10,970 | 5,390 (49) | 3,450 | 1,940 (56) | 892,440 | 641,850 (72) |
| 40,000-50,000 | 1,070 | 150 (14) | 1,420 | 250 (18) | 6,070 | 3,050 (50) | 2,020 | 1,280 (63) | 457,290 | 326,450 (71) |
| 50,000-100,000 | 510 | 90 (18) | 810 | 180 (22) | 4,340 | 2,230 (51) | 1,630 | 1,110 (68) | 354,940 | 252,200 (71) |
| ≥100,000 | 20 | . | 60 | 30 (42) | 220 | 130 (62) | 150 | 110 (70) | 33,280 | 23,470 (71) |
| Unknown | 30 | . | 70 | 10 (14) | 130 | 50 (35) | 100 | 50 (46) | 2,450 | 1,290 (52) |
| **Education level^2,3^** | | | | | | | | | | |
| Primary | 10,990 | 600 (5) | 10,040 | 1,080 (11) | 5,330 | 1,830 (34) | 1,260 | 350 (28) | 77,090 | 42,450 (55) |
| Lower secondary | 10,010 | 770 (8) | 9,850 | 1,440 (15) | 15,430 | 6,890 (45) | 3,540 | 1,440 (41) | 722,690 | 477,920 (66) |
| Upper secondary | 3,750 | 460 (12) | 4,840 | 800 (17) | 22,230 | 10,400 (47) | 7,140 | 3,600 (50) | 1,722,220 | 1,194,510 (69) |
| Bachelor | 1,070 | 190 (18) | 1,440 | 350 (24) | 7,670 | 4,060 (53) | 3,080 | 1,950 (63) | 647,300 | 477,650 (74) |
| Master, doctor | 500 | 120 (24) | 650 | 210 (32) | 2,790 | 1,570 (56) | 1,270 | 890 (70) | 191,240 | 145,280 (76) |
| Unknown | 220 | 60 (25) | 370 | 120 (33) | 1,300 | 650 (50) | 620 | 320 (52) | 74,340 | 48,050 (65) |
| **Medical risk** | | | | | | | | | | |
| Low | 10,080 | 770 (8) | 10,210 | 1,470 (14) | 23,770 | 10,710 (45) | 8,950 | 4,560 (51) | 1,928,250 | 1,320,260 (68) |
| Moderate | 15,380 | 1,300 (8) | 15,840 | 2,330 (15) | 28,100 | 13,340 (47) | 7,060 | 3,540 (50) | 1,354,440 | 957,560 (71) |
| High | 1,090 | 120 (11) | 1,140 | 190 (17) | 2,880 | 1,360 (47) | 890 | 450 (51) | 152,200 | 108,030 (71) |
| **Urbanisation level^2^** | | | | | | | | | | |
| Not urban | 190 | 50 (29) | 200 | 70 (34) | 1,140 | 650 (57) | 900 | 580 (65) | 652,610 | 430,080 (66) |
| Somewhat urban | 790 | 120 (16) | 900 | 180 (20) | 2,000 | 1,170 (58) | 1,320 | 860 (65) | 633,080 | 442,750 (70) |
| Moderately urban | 2,540 | 260 (10) | 3,230 | 550 (17) | 5,460 | 2,910 (53) | 2,410 | 1,420 (59) | 695,670 | 493,830 (71) |
| Highly urban | 6,770 | 630 (9) | 8,150 | 1,300 (16) | 14,850 | 7,310 (49) | 5,400 | 2,660 (49) | 881,110 | 621,300 (71) |
| Extremely urban | 16,270 | 1,120 (7) | 14,710 | 1,900 (13) | 31,300 | 13,370 (43) | 6,870 | 3,030 (44) | 571,370 | 397,240 (70) |
| **Vehicle ownership^2^** | | | | | | | | | | |
| Yes | 12,730 | 1,210 (10) | 14,270 | 2,230 (16) | 32,440 | 16,140 (50) | 10,440 | 5,980 (57) | 2,792,800 | 1,975,180 (71) |
| No | 13,820 | 980 (7) | 12,910 | 1,770 (14) | 22,310 | 9,270 (42) | 6,460 | 2,570 (40) | 642,090 | 410,680 (64) |
| **Distance to vaccination location^2^** | | | | | | | | | | |
| <1km | 2,650 | 210 (8) | 2,500 | 380 (15) | 4,860 | 2,230 (46) | 1,360 | 630 (46) | 158,250 | 112,870 (71) |
| 1-3km | 10,600 | 850 (8) | 9,510 | 1,420 (15) | 22,370 | 10,080 (45) | 5,850 | 2,780 (48) | 744,820 | 524,910 (70) |
| 3-5km | 8,010 | 560 (7) | 7,860 | 1,120 (14) | 17,470 | 7,820 (45) | 4,440 | 2,180 (49) | 671,070 | 468,250 (70) |
| 5-10km | 2,780 | 310 (11) | 4,180 | 610 (15) | 7,170 | 3,720 (52) | 3,250 | 1,870 (58) | 1,020,530 | 703,410 (69) |
| ≥10km | 2,500 | 260 (10) | 3,130 | 470 (15) | 2,870 | 1,560 (54) | 2,000 | 1,090 (55) | 839,150 | 575,760 (69) |

*Note.* Following CBS publication guidelines, all tables exclude frequencies below ten and all numbers and percentages are rounded to the nearest ten to avoid personally identifiable information. Excluded numbers are replaced by “.”.

^1^ Born NL, 1p = Born in the Netherlands with one parent born abroad; Born NL, 2p = Born in the Netherlands with two parents born abroad; Migrant, 1p = Born abroad with one parent born abroad; Migrant, 2p = Born abroad with two parents born abroad; Migrant, 2p NL = Born abroad with two parents born in the Netherlands.

^2^ Measured at household level.

^3^ Education levels are described in more detail in Supplement 2.

Supplement 5: Effect of being a migrant with two foreign-born parents across models

Table S5.1 Associations between being a migrant with two foreign-born parents (reference: born in the Netherlands with one parent born abroad) and being unvaccinated against COVID-19 in the primary vaccination round among persons of Dutch-Caribbean, Moroccan, Surinamese and Turkish origin aged 18 years and older.

| **Country of origin** | **Model 1^1^** | | **Model 2^1^** | | **Model 3^1^** | | **Model 4^1^** | | **Model 5^1^** | | **Model 6^1^** | |
| --- | --- | --- | --- | --- | --- | --- | --- | --- | --- | --- | --- | --- |
|  | aOR | 95% CI | aOR | 95% CI | aOR | 95% CI | aOR | 95% CI | aOR | 95% CI | aOR | 95% CI |
| Moroccan | **2.16** | 2.07-2.26 | **1.80** | 1.72-1.88 | **1.78** | 1.70-1.86 | **1.72** | 1.65-1.80 | **1.71** | 1.64-1.79 | **1.70** | 1.63-1.78 |
| Turkish | **1.10** | 1.06-1.14 | **0.93** | 0.90-0.97 | **0.92** | 0.89-0.96 | **0.90** | 0.87-0.93 | **0.90** | 0.87-0.93 | **0.90** | 0.87-0.94 |
| Surinamese | **1.21** | 1.18-1.25 | **1.09** | 1.06-1.12 | **1.09** | 1.06-1.12 | **1.07** | 1.04-1.10 | **1.07** | 1.04-1.10 | **1.06** | 1.03-1.09 |
| Dutch-Caribbean | **3.39** | 3.28-3.50 | **2.51** | 2.42-2.60 | **2.52** | 2.43-2.61 | **2.48** | 2.40-2.57 | **2.50** | 2.41-2.59 | **2.48** | 2.39-2.57 |

^1^ Model 1: Adjusted for age and sex. Model 2: Model 1 + socioeconomic position, household income and household education level. Model 3: Model 2 + medical risk group. Model 4: Model 3 + urbanisation level of the household. Model 5: Model 4 + household car ownership. Model 6: Model 5 + distance to nearest long-term vaccination location.

Table S5.2 Associations between being a migrant with two foreign-born parents (reference: born in the Netherlands with one parent born abroad) and being unvaccinated in the autumn booster 2022 round among persons of Dutch-Caribbean and Surinamese origin aged 60 years and older.

| **Country of origin** | **Model 1^1^** | | **Model 2^1^** | | **Model 3^1^** | | **Model 4^1^** | | **Model 5^1^** | | **Model 6^1^** | |
| --- | --- | --- | --- | --- | --- | --- | --- | --- | --- | --- | --- | --- |
|  | aOR | 95% CI | aOR | 95% CI | aOR | 95% CI | aOR | 95% CI | aOR | 95% CI | aOR | 95% CI |
| Surinamese | **2.22** | 2.02-2.44 | **1.99** | 1.81-2.19 | **2.02** | 1.83-2.22 | **1.87** | 1.70-2.07 | **1.86** | 1.69-2.05 | **1.84** | 1.67-2.04 |
| Dutch-Caribbean | **2.51** | 2.12-2.99 | **1.99** | 1.66-2.39 | **2.00** | 1.68-2.40 | **1.90** | 1.59-2.29 | **1.88** | 1.57-2.26 | **1.88** | 1.57-2.26 |

^1^ Model 1: Adjusted for age and sex. Model 2: Model 1 + socioeconomic position, household income and household education level. Model 3: Model 2 + medical risk group. Model 4: Model 3 + urbanisation level of the household. Model 5: Model 4 + household car ownership. Model 6: Model 5 + distance to nearest long-term vaccination location.

Supplement 6: Results of the analysis of being unvaccinated the primary vaccination round among persons aged 60 and older

**Descriptive analyses**

The study populations aged ≥60 years included 23,140 persons of Dutch-Caribbean origin, 40,480 of Moroccan origin, 68,060 of Surinamese origin, 39,680 of Turkish origin and 3,890,260 of Dutch origin. The populations of Moroccan and Turkish origin were limited to migrants with two foreign-born parents (Moroccan: 40,400 persons, Turkish: 39,550 persons), because too few individuals had other migration backgrounds. 91.6% of the population aged ≥60 of Dutch origin had received at least one COVID-19 vaccination in the primary vaccination round, compared to 73.4%, 64.9%, 80.8% and 68.1% of the populations of Dutch-Caribbean, Moroccan, Surinamese and Turkish origin, respectively.

**Associations between potential determinants and not having received vaccination**

Similar to the analyses of the 2022 autumn booster round, migration background was removed and the highest income levels were merged in the models for the populations of Moroccan and Turkish origin. Being a student was merged with unknown/other socioeconomic position in all models.

In all populations of non-Dutch origin aged ≥60, female sex was weakly associated with higher odds of not being vaccinated in the primary vaccination round (aORs between 1.13 and 1.24) (Table S6.1). Odds of not being vaccinated also increased with decreasing medical risk (aORs between 0.97 and 1.64). Compared to working in employment, all socioeconomic positions were associated with lower uptake, except being a pensioner or on benefits in the population of Dutch-Caribbean origin. The effects of socioeconomic position were notably strong among persons of Turkish origin. In contrast, younger age, vehicle ownership and further distance to the nearest vaccination location were associated with higher uptake. The effect of vehicle ownership was strong in the population of Surinamese origin (aOR=0.64). Most associations were similar among persons of Dutch origin.

The effects of income and education level differed between the countries of origin. Among persons of Dutch-Caribbean, Surinamese and Dutch origin, odds of being unvaccinated increased strongly as income decreased for incomes below €30,000 (aORs between 1.21 and 10.18). In the populations of Moroccan and Turkish origin, income levels between €10,000 and €40,000 were associated with higher uptake (aORs between 0.61 and 0.85) and only income <10,000 was associated with lower uptake (aOR=1.72 and aOR=2.01, respectively), compared to the highest income level. In the populations of Dutch-Caribbean and Moroccan origin, (lower) secondary and primary education level were associated with lower uptake (aORs between 1.27 and 1.61). Among persons of Dutch origin, odds of being unvaccinated increased as education level decreased. These associations among persons aged ≥60 years differed from those observed for the 2022 autumn booster round.

The effect of migration background could only be estimated in the populations of Dutch-Caribbean and Surinamese origin. Being a migrant with two foreign-born parents was associated with lower uptake (aOR=3.60 and aOR=1.74, respectively). For persons of Dutch-Caribbean origin, adding socioeconomic position, income and education level to the model reduced this aOR, indicating that the effect is mediated by socioeconomic status (Table S6.2). The same was observed for the population of Surinamese origin, although the difference was smaller. Persons of Dutch-Caribbean origin born in the Netherlands with two foreign-born parents (aOR=1.69) and Surinamese migrants with one parent born abroad (aOR=1.44) also had higher odds of being unvaccinated.

**Population attributable fractions of determinants**

The only variable that contributed strongly to being unvaccinated against COVID-19 in the primary vaccination round shared by all four populations aged ≥60 years of non-Dutch origin was not belonging to a medical risk group (PAFs between 9.0% and 18.6%) (Figure S6.1). This is in contrast to the low PAFs for no medical risk in the 2022 autumn booster round. Multiple high PAFs were observed, but these varied between the populations of non-Dutch origin. Income below €30,000 contributed very strongly to being unvaccinated in the ≥60-year-old populations of Dutch-Caribbean and, to a lesser extent, Surinamese origin (PAF=31.1% and PAF=15.9%, respectively). This was not observed for the populations of Moroccan and Turkish origin, because there was little difference and increasing ORs among the income categories above €10,000 in these populations. Like for the 2022 autumn booster round, the PAF of being a migrant and having two foreign-born parents was very high in the populations of Dutch-Caribbean and Surinamese origin (PAF=56.2% and PAF=28.9%, respectively) and could not be assessed for the other populations. The PAFs of being a pensioner and living in an highly or extremely urban area, which had the largest PAFs in the 2022 autumn booster round, were quite large in all populations of non-Dutch origin apart from those of Dutch-Caribbean origin. In the population of Dutch-Caribbean origin, age 60-69 years had a high PAF. Consistent with the regression analyses results, not owning a vehicle had a high PAF among persons of Surinamese origin. Low education level contributed strongly to not being vaccinated in the primary vaccination round in the elderly populations of Dutch-Caribbean and Moroccan origin. Finally, among persons of Turkish origin the PAF of being on benefits was large. All of the aforementioned high PAFs, except being on benefits or a pensioner and living in an highly or extremely urban area, were also observed among persons of Dutch origin.

Table S6.1 Results of multilevel multivariable models of the association between determinants and being unvaccinated in the primary COVID-19 vaccination round among persons of Dutch-Caribbean, Moroccan, Turkish and Dutch origin aged 60 years and older.

|  | **Moroccan origin^1^** | | **Turkish origin^1^** | | **Surinamese origin** | | **Dutch-Caribbean origin** | | **Dutch origin** | |
| --- | --- | --- | --- | --- | --- | --- | --- | --- | --- | --- |
|  | **aOR** | **95% CI** | **aOR** | **95% CI** | **aOR** | **95% CI** | **aOR** | **95% CI** | **aOR** | **95% CI** |
| **Sex^2^** | | | | | | | | | | |
| Male | (ref) |  |  |  | (ref) |  | (ref) |  | (ref) |  |
| Female | **1.21** | 1.16-1.26 | **1.13** | 1.08-1.18 | **1.16** | 1.12-1.21 | **1.24** | 1.16-1.31 | **1.09** | 1.09-1.10 |
| **Age^3^** | | | | | | | | | | |
| ≥80 | (ref) |  | (ref) |  | (ref) |  | (ref) |  | (ref) |  |
| 70-79 | **0.75** | 0.71-0.80 | 1.01 | 0.95-1.09 | **0.80** | 0.75-0.86 | **0.86** | 0.76-0.98 | **0.64** | 0.63-0.65 |
| 60-70 | **0.75** | 0.71-0.80 | **0.92** | 0.86-0.99 | **0.85** | 0.80-0.91 | **1.22** | 1.09-1.38 | **0.77** | 0.76-0.77 |
| **Migration background^4^** | | | | | | | | | | |
| Born NL, 1p | NA | | NA | | (ref) |  | (ref) |  | NA | |
| Born NL, 2p |  |  |  |  | 1.26 | 0.94-1.67 | **1.69** | 0.86-3.14 |  |  |
| Migrant, 1p |  |  |  |  | **1.44** | 1.14-1.81 | 1.07 | 0.80-1.45 |  |  |
| Migrant, 2p |  |  |  |  | **1.74** | 1.54-1.98 | **3.60** | 2.85-4.60 |  |  |
| Migrant, 2p NL |  |  |  |  | 0.85 | 0.61-1.16 | **0.69** | 0.53-0.91 |  |  |
| **Socioeconomic position^5^** | | | | | | | | | | |
| Employee | (ref) |  | (ref) |  | (ref) |  | (ref) |  | (ref) |  |
| Self-employed | **1.51** | 1.21-1.88 | **1.53** | 1.27-1.83 | **1.27** | 1.11-1.45 | **1.33** | 1.08-1.63 | **1.69** | 1.65-1.72 |
| Pensioner | **1.88** | 1.71-2.08 | **2.82** | 2.53-3.13 | **1.12** | 1.05-1.20 | **0.86** | 0.78-0.95 | **0.94** | 0.92-0.95 |
| On benefits | **1.84** | 1.67-2.03 | **2.49** | 2.24-2.77 | **1.39** | 1.30-1.50 | 1.11 | 0.99-1.24 | **1.43** | 1.40-1.45 |
| Other/unknown | **2.04** | 1.79-2.32 | **3.28** | 2.85-3.77 | **1.34** | 1.15-1.56 | **1.30** | 1.04-1.62 | **1.13** | 1.11-1.16 |
| **Income^6^** | | | | | | | | | | |
| ≥100,000 | (ref) |  | (ref) |  | (ref) |  | (ref) |  | (ref) |  |
| 50,000-100,000 |  |  |  |  | 1.14 | 0.78-1.72 | 1.32 | 0.75-2.51 | 0.99 | 0.94-1.04 |
| 40,000-50,000 | 0.83 | 0.68-1.02 | 0.86 | 0.72-1.02 | 1.37 | 0.94-2.05 | 1.72 | 0.99-3.24 | **1.06** | 1.01-1.11 |
| 30,000-40,000 | **0.83** | 0.69-0.99 | **0.85** | 0.73-1.00 | **1.51** | 1.05-2.27 | **1.97** | 1.14-3.70 | **1.21** | 1.15-1.26 |
| 20,000-30,000 | **0.69** | 0.58-0.82 | **0.75** | 0.64-0.87 | **1.82** | 1.26-2.73 | **2.76** | 1.60-5.18 | **1.56** | 1.49-1.64 |
| 10,000-20,000 | **0.61** | 0.52-0.73 | **0.70** | 0.60-0.87 | **2.33** | 1.62-3.50 | **4.13** | 2.39-7.75 | **2.63** | 2.51-2.76 |
| <10,000 | **1.72** | 1.36-2.18 | **2.01** | 1.65-2.44 | **7.78** | 5.30-11.80 | **6.78** | 3.88-12.89 | **10.18** | 9.69-10.71 |
| **Education level^7^** | | | | | | | | | | |
| Master, doctor | (ref) |  | (ref) |  | (ref) |  | (ref) |  | (ref) |  |
| Bachelor | 1.05 | 0.87-1.29 | 1.07 | 0.91-1.28 | 0.99 | 0.88-1.10 | 1.10 | 0.92-1.32 | **1.14** | 1.12-1.17 |
| Upper secondary | 1.19 | 1.00-1.42 | 1.09 | 0.94-1.28 | 0.95 | 0.86-1.06 | **1.27** | 1.08-1.50 | **1.21** | 1.18-1.23 |
| Lower secondary | **1.28** | 1.08-1.53 | 0.99 | 0.85-1.15 | 1.00 | 0.90-1.11 | **1.31** | 1.10-1.57 | **1.41** | 1.38-1.44 |
| Primary | **1.40** | 1.18-1.67 | 1.00 | 0.86-1.17 | 0.92 | 0.82-1.03 | **1.61** | 1.33-1.95 | **1.92** | 1.86-1.97 |
| **Medical risk^8^** | | | | | | | | | | |
| High risk | (ref) |  | (ref) |  | (ref) |  | (ref) |  | (ref) |  |
| Moderate risk | **1.24** | 1.10-1.39 | **1.23** | 1.10-1.39 | 0.97 | 0.87-1.07 | **1.25** | 1.07-1.47 | **0.83** | 0.81-0.84 |
| None | **1.46** | 1.30-1.64 | **1.59** | 1.41-1.79 | **1.64** | 1.49-1.81 | **1.64** | 1.41-1.92 | **1.02** | 1.00-1.04 |
| **Urbanisation level^9^** | | | | | | | | | | |
| Not urban | (ref) |  | (ref) |  | (ref) |  | (ref) |  | (ref) |  |
| Somewhat urban | 0.97 | 0.71-1.32 | 0.79 | 0.58-1.09 | 0.86 | 0.70-1.04 | 0.84 | 0.67-1.05 | **0.83** | 0.83-0.85 |
| Moderately  urban | 0.98 | 0.73-1.32 | 0.83 | 0.62-1.11 | 0.85 | 0.72-1.02 | 0.97 | 0.80-1.18 | **0.82** | 0.81-0.83 |
| Highly urban | 1.07 | 0.81-1.43 | 0.93 | 0.70-1.25 | 1.06 | 0.90-1.25 | 1.04 | 0.87-1.25 | **0.88** | 0.87-0.89 |
| Extremely urban | 1.27 | 0.96-1.69 | 1.21 | 0.92-1.62 | **1.23** | 1.04-1.47 | 1.10 | 0.92-1.32 | 0.99 | 0.98-1.00 |
| **Vehicle ownership^10^** | | | | | | | | | | |
| No | (ref) |  | (ref) |  | (ref) |  | (ref) |  | (ref) |  |
| Yes | **0.89** | 0.85-0.94 | **0.94** | 0.89-0.98 | **0.64** | 0.61-0.67 | **0.89** | 0.82-0.96 | **0.62** | 0.61-0.63 |
| **Distance to vaccination location^11^** | | | | | | | | | | |
| <1km | (ref) |  | (ref) |  | (ref) |  | (ref) |  | (ref) |  |
| 1-3km | 0.96 | 0.89-1.02 | **0.84** | 0.78-0.90 | **0.94** | 0.88-1.00 | 0.94 | 0.84-1.05 | **0.97** | 0.95-0.99 |
| 3-5km | 0.97 | 0.90-1.04 | **0.76** | 0.71-0.82 | **0.87** | 0.82-0.93 | **0.86** | 0.76-0.96 | **0.97** | 0.95-0.99 |
| 5-10km | **0.90** | 0.83-0.98 | **0.86** | 0.79-0.93 | **0.81** | 0.75-0.88 | **0.84** | 0.74-0.95 | **0.94** | 0.92-0.96 |
| ≥10km | **0.87** | 0.80-0.95 | **0.76** | 0.69-0.82 | **0.81** | 0.74-0.89 | **0.81** | 0.71-0.92 | 1.00 | 0.97-1.02 |

^1^ Population limited to migrants with two parents born abroad.

^2^ Adjusted for age and migration background (populations of Dutch-Caribbean and Surinamese origin only).

^3^ Adjusted for sex and migration background (populations of Dutch-Caribbean and Surinamese origin only).

^4^ Adjusted for age and sex. Born NL, 1p = Born in the Netherlands with one parent born abroad; Born NL, 2p = Born in the Netherlands with two parents born abroad; Migrant, 1p = Born abroad with one parent born abroad; Migrant, 2p = Born abroad with two parents born abroad; Migrant, 2p NL = Born abroad with two parents born in the Netherlands. Not applicable (N.A.) for the populations of Moroccan, Turkish and Dutch origin.

^5^ Adjusted for age, sex, migration background (populations of Dutch-Caribbean and Surinamese origin only), income and education level.

^6^ Measured at household level. Adjusted for age, sex, migration background (populations of Dutch-Caribbean and Surinamese origin only), socioeconomic position and education level.

^7^ Measured at household level. Adjusted for age, sex, migration background (populations of Dutch-Caribbean and Surinamese origin only), socioeconomic position and income. Education levels are described in more detail in Supplement 2.

^8^ Adjusted for age, sex, migration background (populations of Dutch-Caribbean and Surinamese origin only), socioeconomic position, income and education level.

^9^ Measured at household level. Adjusted for age, sex, migration background (populations of Dutch-Caribbean and Surinamese origin only), socioeconomic position, income, education level and medical risk.

^10^ Measured at household level. Adjusted for age, sex, migration background (populations of Dutch-Caribbean and Surinamese origin only), socioeconomic position, income, education level, medical risk and urbanisation level.

^11^ Measured at household level. Adjusted for age, sex, migration background (populations of Dutch-Caribbean and Surinamese origin only), socioeconomic position, income, education level, medical risk, urbanisation level and vehicle ownership.

Table S6.2 Associations between being a migrant with two foreign-born parents (reference: born in the Netherlands with one parent born abroad) and not being unvaccinated in the primary vaccination round among persons of Dutch-Caribbean and Surinamese origin aged 60 years and older.

| **Country of origin** | **Model 1^1^** | | **Model 2^1^** | | **Model 3^1^** | | **Model 4^1^** | | **Model 5^1^** | | **Model 6^1^** | |
| --- | --- | --- | --- | --- | --- | --- | --- | --- | --- | --- | --- | --- |
|  | aOR | 95% CI | aOR | 95% CI | aOR | 95% CI | aOR | 95% CI | aOR | 95% CI | aOR | 95% CI |
| Surinamese | **1.74** | 1.54-1.98 | **1.50** | 1.32-1.70 | **1.61** | 1.42-1.83 | **1.52** | 1.34-1.73 | **1.49** | 1.31-1.70 | **1.48** | 1.30-1.68 |
| Dutch-Caribbean | **3.60** | 2.85-4.60 | **2.76** | 2.17-3.56 | **2.78** | 2.19-3.59 | **2.71** | 2.13-3.50 | **2.68** | 2.10-3.46 | **2.68** | 2.10-3.47 |

^1^ Model 1: Adjusted for age and sex. Model 2: Model 1 + socioeconomic position, household income and household education level. Model 3: Model 2 + medical risk group. Model 4: Model 3 + urbanisation level of the household. Model 5: Model 4 + household car ownership. Model 6: Model 5 + distance to nearest long-term vaccination location.

Figure S6.1 Population attributable fractions of the contribution of each determinant to being unvaccinated in the primary vaccination round by country of origin among persons aged 60 years and older.

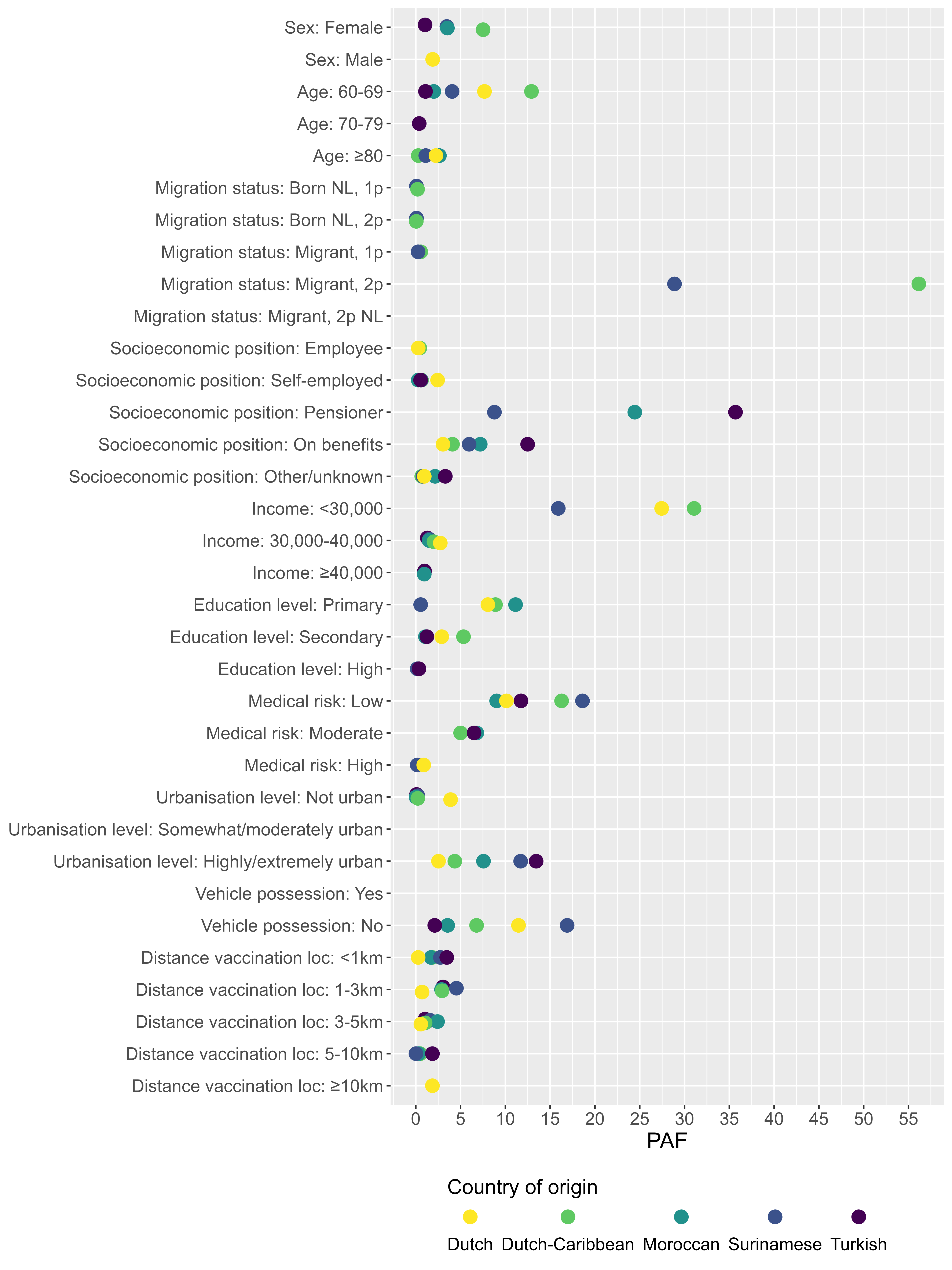

^1^ Populations of Moroccan and Turkish origin were limited to migrants with two foreign-born parents.

^2^ Born NL, 1p = Born in the Netherlands with one parent born abroad; Born NL, 2p = Born in the Netherlands with two parents born abroad; Migrant, 1p = Born abroad with one parent born abroad; Migrant, 2p = Born abroad with two parents born abroad; Migrant, 2p NL = Born abroad with two parents born in the Netherlands. Not applicable for Moroccan, Turkish and Dutch country of origin.

^3^ Income, education level, urbanisation level, vehicle possession and distance to vaccination location were measured at household level.

^4^ The reference category for each country of origin does not feature a dot for that country of origin.
